# Drug utilization across occupations among pregnant women working in the healthcare and social assistance sector in France: a nationwide population-based study

**DOI:** 10.64898/2026.01.12.26343952

**Authors:** Mélanie Araujo, Justine Bénévent, Christine Damase-Michel, Yolande Esquirol, Anna-Belle Beau

## Abstract

**Purpose:** Medication use during pregnancy is common, but patterns may differ across occupational subgroups, particularly in healthcare and social assistance where working conditions, education, and health literacy vary. We aimed to compare medications dispensed to pregnant women across occupations within this sector.

**Methods:** We conducted a nationwide population-based study using the French EDP-Santé database, linking occupational and socioeconomic data with healthcare reimbursement records. Pregnancies from 2009–2015 among women aged 15–55 years were identified. Medication dispensing during the year before and during pregnancy was described by occupational groups using the Anatomical Therapeutic Chemical classification. Comparisons were performed within healthcare occupations.

**Results:** Among 321,569 pregnancies, 38,860 (12%) involved women in healthcare and social assistance. Before pregnancy, 91–95% received at least one medication (mean: 8–12). Psychotropic medication (including antidepressants and benzodiazepines) dispensing was higher among hospital service workers (18% [95% confidence interval (CI):17–19]), healthcare assistants (16% [95% CI:15–16]), and social workers (14% [95% CI:13–15]) than among physicians (11% [95% CI:10–13]), rehabilitation professionals (10% [95% CI:8–12]), and nurses (10% [95% CI:9–10]). Similar patterns were observed for analgesic and musculoskeletal medications. During pregnancy, differences were less pronounced. Folic acid, recommended before and early in pregnancy, was lower among hospital service workers (30% [95% CI:29–32]) than other healthcare workers (39–53%).

**Conclusion:** Medication dispensing is common among pregnant healthcare workers, with marked variation across occupations. These differences may reflect disparities in working conditions or socioeconomic level, underlining the need for tailored maternal health support in specific occupational groups.

## Introduction

In France, over 90% of pregnant women receive at least one medication during pregnancy, mainly for routine prenatal supplements (iron and folic acid), acute conditions (digestive disorders or urinary tract infections) or chronic conditions ^1^. While this highlights the overall high exposure to medications in pregnancy, it likely masks important differences between population subgroups. Certain groups may be particularly vulnerable to higher or inappropriate medication use due to differences in working conditions, socioeconomic level, and comorbidities.

Women working in the healthcare and social assistance sector represent over 70% of the workforce ^2^, and encompass a wide range of occupations from physicians and midwives to nursing assistants and hospital service workers with varying educational backgrounds, job demands, and work-related stressors. These factors may influence both health status and patterns of medication use during pregnancy.

High-quality prenatal care is a key indicator of the overall quality of healthcare. It relies on regular follow-up visits, screening, and appropriate use of medication, such as timely administration of folic acid and vaccines, and the avoidance of potentially harmful drugs. Inadequate prenatal follow-up or inappropriate medication exposure can have serious consequences for both the mother and the unborn child. In France, little is known about the prenatal care and medication use of pregnant healthcare professionals. Evidence from other settings suggests higher psychoactive substance use among healthcare professionals. For instance, a U.S. study reported signs of alcohol dependence in 15% of physicians versus 9% in the general population ^3^. A 2021 French survey highlighted inadequate care for depression among healthcare professionals, resulting in higher use of anxiolytics and hypnotics ^4^. However, broader data on prescription and dispensing of medications (including psychotropics, analgesics, and non-steroidal anti-inflammatory drugs (NSAIDs) during pregnancy are lacking. Moreover, recent and alarming reports on the working conditions of healthcare workers in France highlight the need to better understand the health of pregnant healthcare professionals ^5^.

This study aimed at evaluating medication dispensing before and during pregnancy, among women working in the healthcare and social assistance sector, comparing intra- and inter-sector occupations. By comparing different occupations within this sector and with pregnant women employed in other sectors, this study examined the impact of occupational sectors on medication patterns.

## Material and methods

### Data source

We utilized data from the EDP-Santé 2017, a French administrative database linking the Permanent Demographic Sample (EDP) from the French National Institute for Statistics and Economic Studies (INSEE) with the National Health Data System (SNDS) ^6^. The EDP covers approximately 4% of the French population and combines detailed socioeconomic data and occupational information with comprehensive outpatient (DCIR) and inpatient (PMSI) healthcare data, including medication dispensing, medical procedures and hospital diagnoses). Additional details are provided in the online supplements **(S1 Method)**.

### Pregnancy identification

Pregnancies occurring between January 2009 and December 2015 were identified using hospital discharge diagnoses and medical procedures recorded in the SNDS. The start of pregnancy was defined as the first day of the last menstrual period (LMP), estimated primarily from gestational age recorded during hospital stays, mainly based on the first-trimester ultrasound. When gestational age was missing, the median gestational age by pregnancy outcome observed in the general population was assigned, as done previously ^7^. Full algorithms and codes are detailed in the online supplements **(S1 Table)**.

### Study population

We included women aged 15 to 55 years at the start of pregnancy. The study population was divided into two groups: (1) healthcare and social assistance professionals; and (2) non-healthcare professionals.

### Occupational classification

Occupation status during the periconceptional period (defined as the year of the LMP date) was mainly derived from employer declarations recorded in the EDP “active worker panel”, supplemented by census data. Occupations were identified using the level-four French occupational classification codes. The healthcare and social assistance professionals were grouped into eight occupational groups: (1) physicians: medical physicians, dental surgeons and midwives; (2) psychologists; (3) nurses; (4) social workers; (5) rehabilitation professionals; (6) healthcare assistants: nursing, dental or medical or psychologist assistants, and, childcare auxiliaries; (7) hospital service workers; and (8) others healthcare professionals, not classified elsewhere. assistants and hospital service workers). Occupations were subsequently aggregated into three socioeconomic occupational groups. Non-healthcare professionals were selected from comparable socioeconomic levels. We also retrieved data on sector of employment (healthcare, social assistance and public administration) and employer type (public and private). Classification details are provided in **S2 Table.**

### Medication exposure

Medication dispensing data were obtained from the SNDS and classified using the Anatomical, Therapeutic and Chemical (ATC) system. We examined all medications dispensed during the year preceding and throughout pregnancy. Medications were first described at the ATC level 1, followed by analyses of selected classes of interest, including folic acid, psychotropic medications (benzodiazepines and antidepressants), analgesics, musculo-skeletal medications, and NSAIDs. ATC codes are listed in **S3 Table**.

### Statistical analysis

The unit of analysis was the pregnancy episode. Continuous variables were described using means and standard deviation (SD), and categorical variables using percentages with 95% confidence interval (CI). We compared medication use across the occupational groups (inter-sector comparisons) and with non-healthcare professionals (intra-sector comparisons), stratified by socio-economic level. Differences were considered statistically significant when there was no overlap between the 95% CIs. This conservative approach was chosen due to the multiple comparison groups and the challenge to define a single relevant reference group. Analyses complied with French data protection regulations, and results based on fewer than 10 individuals were not reported.

Data management and creation of study variables were performed using the framework Apache Spark® in RStudio® version 4.3.3 environment of the SparklyR package, facilitating the manipulation of massive data. Formal analyses were performed using SAS Enterprise Guide® version 8.3 (SAS Institute Inc., Cary, NC, USA) and RStudio® version 4.3.3. All analyses were carried out in a secure environment provided by the CASD (Centre d’Accès Sécurisé aux Données (Ref. 10.34724/CASD)).

## Results

### Characteristics of the study population

Among the 3,732,088 individuals in the EDP-Santé, we identified 321,569 pregnancy episodes from 214,423 women aged 15 to 55 years with pregnancy onset between January 2009 and December 2015 (**Figure 1**). Of these, 38,860 pregnancies (12%) occurred among women employed in the healthcare and social assistance sector, and 164,384 pregnancies among women employed in other sectors with comparable socioeconomic levels.

**Figure 1.** Flowchart describing the study population selection. A pregnancy can be identified in both the active worker panel and the annual census surveys, but was counted once in the dataset.

Within the healthcare and social assistance sector, 8% were executives, 48% intermediate occupations and 44% employees, compared with 15%, 20% and 65% respectively in other sectors. The healthcare and social assistance population included 11,004 (28%) healthcare assistants, 9,521 (25%) nurses, 5,602 (14%) hospital service workers, 4,640 (12%) social workers, 2,212 (6%) physicians, 959 (3%) rehabilitation professionals, 817 (2%) psychologists, and 4,105 (11%) others healthcare professionals such as pharmacists, medical technicians and opticians. Mean maternal age at pregnancy onset was 30.0 ± 5.0 years, ranging from 28.2 ± 6.1 years for hospital service workers to 32.3 ± 4.0 for psychologists (**Table 1**). Employment in the public sector varied across occupations from 35% among social work professionals to 81% among physicians.

**Table 1:** Population characteristics.

|  | Physicians<br>N=2,212 | Psychologists<br>N=817 | Nurses N=9,521 | Social workers<br>N=4,640 | Rehabilitation<br>professionals<br>N=959 | Healthcare<br>assistants<br>N=11,004 | Hospital service<br>workers<br>N=5,602 | Other N=4,105 |
| --- | --- | --- | --- | --- | --- | --- | --- | --- |
| **Age at pregnancy onset (years)** |  |  |  |  |  |  |  |  |
| Mean±SD | 31.3±4.3 | 32.3±4.0 | 30.2±4.5 | 31.0±4.5 | 30.6±4.1 | 29.7±5.2 | 28.2±6.1 | 30.0±4.5 |
| < 25 | 70 (3.2%) | 5 (0.6%) | 749 (7.9%) | 238 (5.1%) | 37 (3.9%) | 1,742 (15.8%) | 1,800 (32.1%) | 369 (9.0%) |
| [25-35[ | 1,655 (74.8%) | 586 (71.7%) | 7,153 (75.1%) | 3,422 (73.8%) | 756 (78.8%) | 7,216 (66.6%) | 2,862 (51.1%) | 3,050 (74.3%) |
| ≥ 35 | 487 (22.0%) | 226 (27.7%) | 1,619 (17.0%) | 980 (21.1%) | 166 (17.3%) | 2,046 (18.6%) | 940 (16.8%) | 686 (16.7%) |
| **Sector of employment<sup>a</sup> (n (%))** |  |  |  |  |  |  |  |  |
| Health | 1,914 (89.7%) | 349 (44.6%) | 7,819 (83.7%) | 323 (7.2%) | 452 (55.7%) | 5,221 (48.2%) | 2,347 (43.2%) | 1,416 (35.8%) |
| Social care | 47 (2.2%) | 299 (38.2%) | 936 (10.0%) | 2,774 (61.6%) | 277 (34.2%) | 4,355 (40.2%) | 2,917 (53.7%) | 40 (1.0%) |
| Public administration | 73 (3.4%) | 94 (12.0%) | 220 (2.4%) | 925 (20.5%) | 17 (2.1%) | 575 (5.3%) | 35 (0.6%) | 37 (0.9%) |
| Others<sup>b</sup> | 100 (4.7%) | 40 (5.1%) | 368 (4.0%) | 480 (10.7%) | 65 (8.0%) | 675 (6.2%) | 134 (2.5%) | 2,466 (62.2%) |
| Missing | 78 | 35 | 178 | 138 | 148 | 178 | 169 | 146 |
| **Employer type (n (%))** |  |  |  |  |  |  |  |  |
| Public | 1,735 (81.3%) | 389 (49.7%) | 6,130 (65.5%) | 1,556 (34.6%) | 324 (40.0%) | 5,086 (46.9%) | 2,599 (47.8%) | 633 (16.0%) |
| Private | 399 (18.7%) | 393 (50.3%) | 3,222 (34.5%) | 2,947 (65.4%) | 487 (60.0%) | 5,748 (53.1%) | 2,841 (52.2%) | 3,330 (84.0%) |
| Missing | 78 | 35 | 169 | 137 | 148 | 170 | 162 | 142 |

**Footnote:** SD, standard deviation; <sup>a</sup> Occupational sectors were categorized using the French A38 classification (derived from NACE Rev.2, European classification of economic
activities); <sup>b</sup>“others” includes for instance : wholesale and retail trade, administrative and support service activities, other service activities, education, or legal, accounting and
management activities.

### Medication dispensing before pregnancy

Overall 91% to 95% of women were dispensed at least one medication in the year preceding pregnancy. The mean number of different medications ranged from 8–9 among physicians, nurses and rehabilitation professionals to 12 among hospital service workers (**Table 2**). In inter-sector comparisons, physicians, nurses and rehabilitation professionals were dispensed significantly fewer medications than non-healthcare workers with similar socioeconomic level, whereas hospital service workers were dispensed significantly more (**Table 2**). Marked differences were observed across therapeutic classes (**Table 3**). Intra-sector comparisons showed that nervous system medications (ATC N) were dispensed more frequently to hospital service workers (77% [95% CI 76–78]), healthcare assistants (75% [95% CI 74–76]), social workers (74% [95% CI 73–76]), and psychologists (68% [95% CI 65–71]) than to rehabilitation professionals (62% [95% CI 59–65]), nurses (61% [95% CI 60–62]) and physicians (60% [95% CI 58–62]). Inter-sector comparisons showed that the latter group was dispensed significantly fewer nervous system medications than non-healthcare workers with similar socioeconomic level, while the former group was dispensed significantly more. For musculoskeletal system medications (ATC M), intra-sector comparisons showed that dispensing was significantly more frequent among hospital service workers (60% [95% CI 59–61]) than among physicians (34% [95% CI 32–36]). A similar pattern was seen for respiratory system medications (ATC R), with higher dispensing among social workers (54% [95% CI 53–56]) compared with physicians (36% [95% CI 34–38]). Conversely, blood and blood-forming organs medications (ATC B) were more significantly more frequently dispensed to physicians (41% [95% CI 39–43]), rehabilitation professionals (40% [95% CI 37–43]) and psychologists (39% (95% CI 36–42)) than to healthcare assistants (33% [95% CI 32–34]), or hospital service workers (27% [95% CI 26v28]). Inter-sector comparisons showed that, compared with non-healthcare workers with similar socioeconomic level, physicians, rehabilitation professionals, healthcare assistants were significantly more likely to be dispensed ATC B medications (**Table 3**).

**Table 2:**
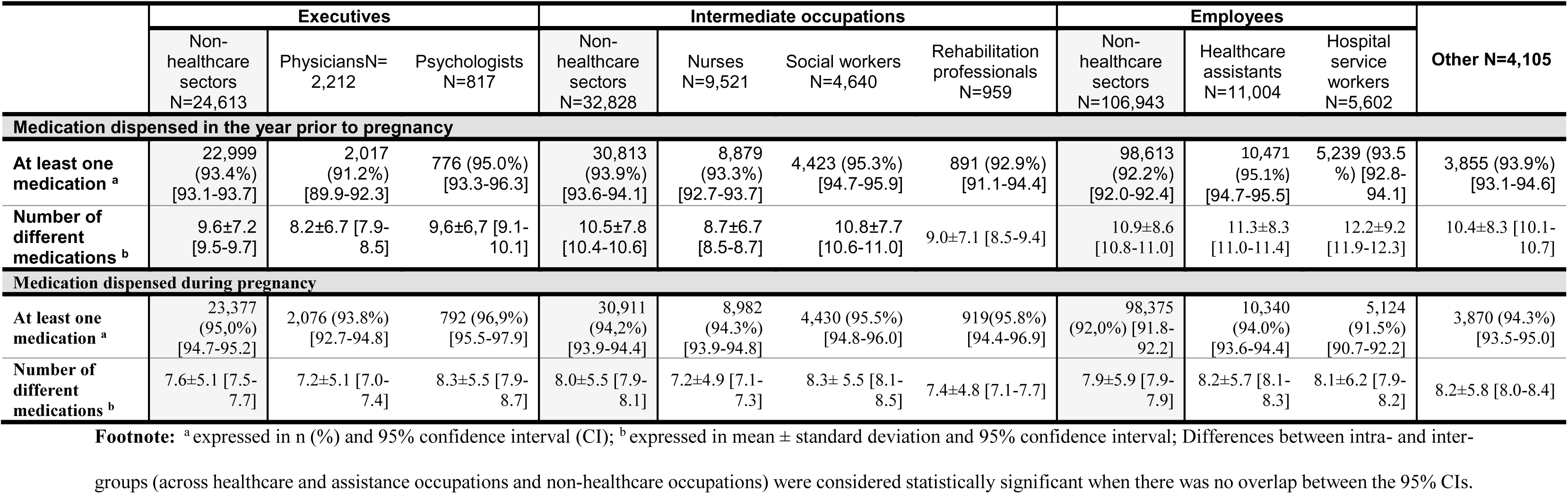
Medication dispensed in the year prior to or during pregnancy by occupational groups.

|  | Executives |  |  | Intermediate occupations |  |  |  | Employees |  |  | Other N=4,105 |
| --- | --- | --- | --- | --- | --- | --- | --- | --- | --- | --- | --- |
|  | Non-healthcare sectors<br>N=24,613 | PhysiciansN=2,212 | Psychologists<br>N=817 | Non-healthcare sectors<br>N=32,828 | Nurses<br>N=9,521 | Social workers<br>N=4,640 | Rehabilitation professionals<br>N=959 | Non-healthcare sectors<br>N=106,943 | Healthcare assistants<br>N=11,004 | Hospital service workers<br>N=5,602 |  |
| Medication dispensed in the year prior to pregnancy |  |  |  |  |  |  |  |  |  |  |  |
| At least one medication <sup>a</sup> | 22,999<br>(93.4%)<br>[93.1-93.7] | 2,017<br>(91.2%)<br>[89.9-92.3] | 776 (95.0%)<br>[93.3-96.3] | 30,813<br>(93.9%)<br>[93.6-94.1] | 8,879<br>(93.3%)<br>[92.7-93.7] | 4,423 (95.3%)<br>[94.7-95.9] | 891 (92.9%)<br>[91.1-94.4] | 98,613<br>(92.2%)<br>[92.0-92.4] | 10,471<br>(95.1%)<br>[94.7-95.5] | 5,239 (93.5%)<br>[92.8-94.1] | 3,855 (93.9%)<br>[93.1-94.6] |
| Number of different medications <sup>b</sup> | 9.6±7.2<br>[9.5-9.7] | 8.2±6.7 [7.9-8.5] | 9.6±6.7 [9.1-10.1] | 10.5±7.8<br>[10.4-10.6] | 8.7±6.7<br>[8.5-8.7] | 10.8±7.7<br>[10.6-11.0] | 9.0±7.1 [8.5-9.4] | 10.9±8.6<br>[10.8-11.0] | 11.3±8.3<br>[11.0-11.4] | 12.2±9.2<br>[11.9-12.3] | 10.4±8.3 [10.1-10.7] |
| Medication dispensed during pregnancy |  |  |  |  |  |  |  |  |  |  |  |
| At least one medication <sup>a</sup> | 23,377<br>(95.0%)<br>[94.7-95.2] | 2,076 (93.8%)<br>[92.7-94.8] | 792 (96.9%)<br>[95.5-97.9] | 30,911<br>(94.2%)<br>[93.9-94.4] | 8,982<br>(94.3%)<br>[93.9-94.8] | 4,430 (95.5%)<br>[94.8-96.0] | 919(95.8%)<br>[94.4-96.9] | 98,375<br>(92.0%) [91.8-92.2] | 10,340<br>(94.0%)<br>[93.6-94.4] | 5,124<br>(91.5%)<br>[90.7-92.2] | 3,870 (94.3%)<br>[93.5-95.0] |
| Number of different medications <sup>b</sup> | 7.6±5.1 [7.5-7.7] | 7.2±5.1 [7.0-7.4] | 8.3±5.5 [7.9-8.7] | 8.0±5.5 [7.9-8.1] | 7.2±4.9 [7.1-7.3] | 8.3± 5.5 [8.1-8.5] | 7.4±4.8 [7.1-7.7] | 7.9±5.9 [7.9-7.9] | 8.2±5.7 [8.1-8.3] | 8.1±6.2 [7.9-8.2] | 8.2±5.8 [8.0-8.4] |

**Footnote:** <sup>a</sup> expressed in n (%) and 95% confidence interval (CI); <sup>b</sup> expressed in mean ± standard deviation and 95% confidence interval; Differences between intra- and inter-groups (across healthcare and assistance occupations and non-healthcare occupations) were considered statistically significant when there was no overlap between the 95% CIs.

**Table 3:**
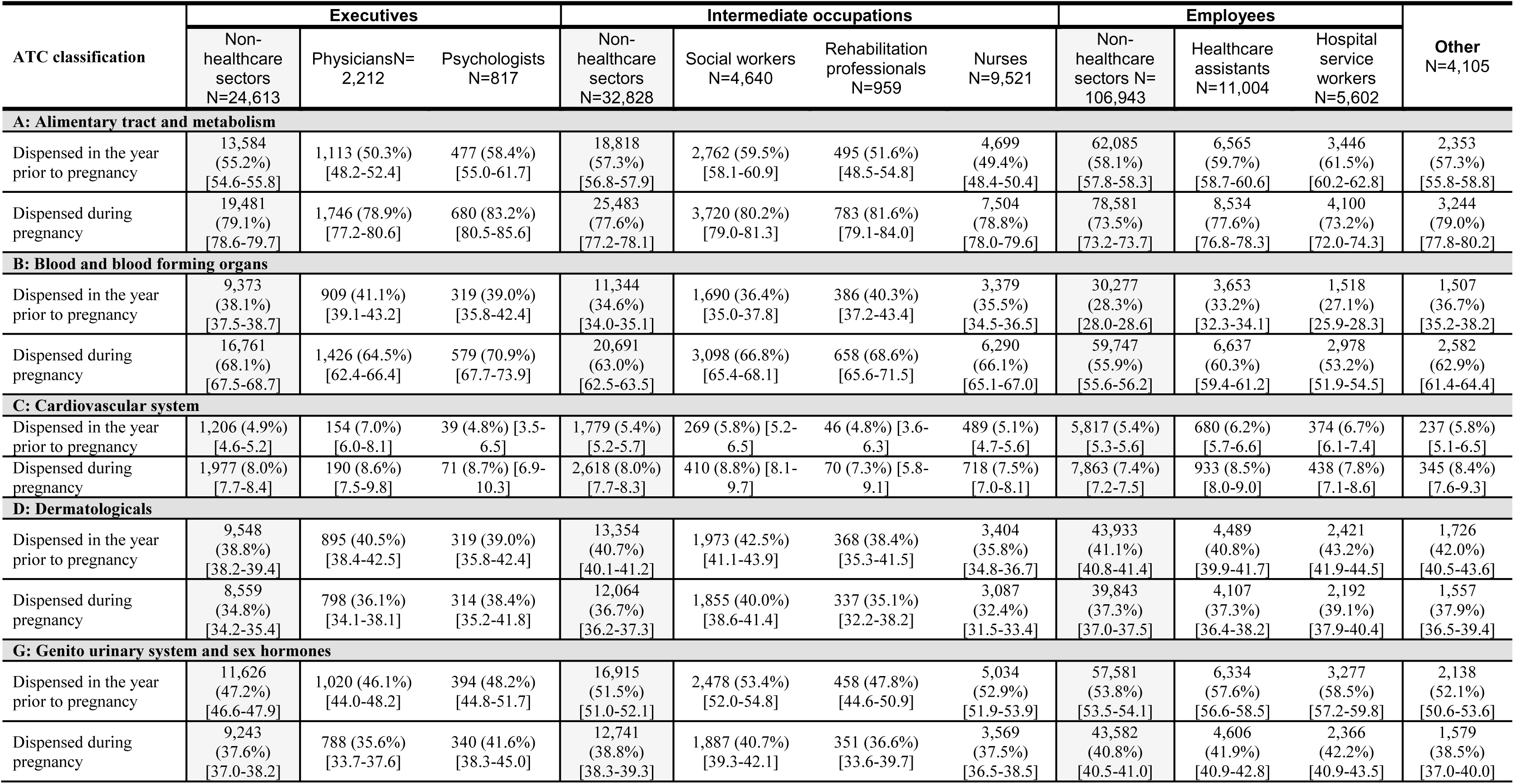
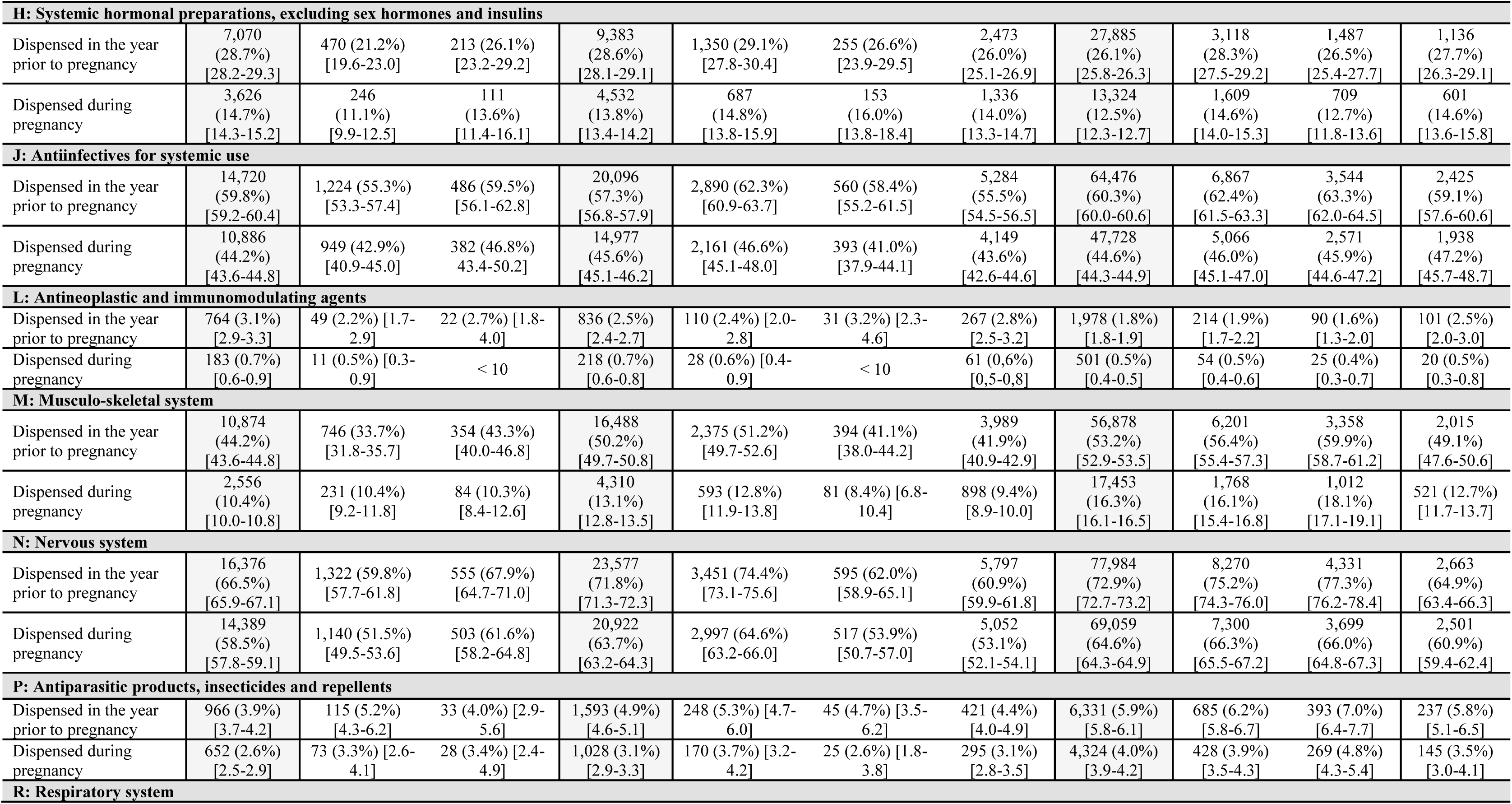
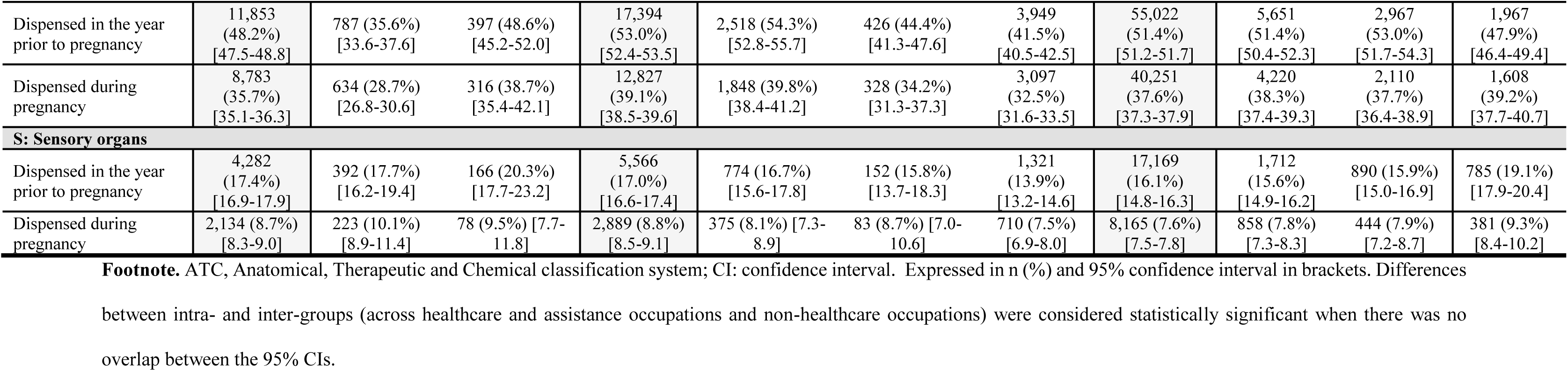
Prevalence of medication dispensed by first level of ATC classification in the year prior to pregnancy and during pregnancy by occupational groups.

| ATC classification | Executives |  |  | Intermediate occupations |  |  |  | Employees |  |  | Other<br>N=4,105 |
| --- | --- | --- | --- | --- | --- | --- | --- | --- | --- | --- | --- |
|  | Non-healthcare sectors<br>N=24,613 | PhysiciansN=2,212 | Psychologists<br>N=817 | Non-healthcare sectors<br>N=32,828 | Social workers<br>N=4,640 | Rehabilitation professionals<br>N=959 | Nurses<br>N=9,521 | Non-healthcare sectors N=106,943 | Healthcare assistants<br>N=11,004 | Hospital service workers<br>N=5,602 |  |
| A: Alimentary tract and metabolism |  |  |  |  |  |  |  |  |  |  |  |
| Dispensed in the year prior to pregnancy | 13,584<br>(55.2%)<br>[54.6-55.8] | 1,113 (50.3%)<br>[48.2-52.4] | 477 (58.4%)<br>[55.0-61.7] | 18,818<br>(57.3%)<br>[56.8-57.9] | 2,762 (59.5%)<br>[58.1-60.9] | 495 (51.6%)<br>[48.5-54.8] | 4,699<br>(49.4%)<br>[48.4-50.4] | 62,085<br>(58.1%)<br>[57.8-58.3] | 6,565<br>(59.7%)<br>[58.7-60.6] | 3,446<br>(61.5%)<br>[60.2-62.8] | 2,353<br>(57.3%)<br>[55.8-58.8] |
| Dispensed during pregnancy | 19,481<br>(79.1%)<br>[78.6-79.7] | 1,746 (78.9%)<br>[77.2-80.6] | 680 (83.2%)<br>[80.5-85.6] | 25,483<br>(77.6%)<br>[77.2-78.1] | 3,720 (80.2%)<br>[79.0-81.3] | 783 (81.6%)<br>[79.1-84.0] | 7,504<br>(78.8%)<br>[78.0-79.6] | 78,581<br>(73.5%)<br>[73.2-73.7] | 8,534<br>(77.6%)<br>[76.8-78.3] | 4,100<br>(73.2%)<br>[72.0-74.3] | 3,244<br>(79.0%)<br>[77.8-80.2] |
| B: Blood and blood forming organs |  |  |  |  |  |  |  |  |  |  |  |
| Dispensed in the year prior to pregnancy | 9,373<br>(38.1%)<br>[37.5-38.7] | 909 (41.1%)<br>[39.1-43.2] | 319 (39.0%)<br>[35.8-42.4] | 11,344<br>(34.6%)<br>[34.0-35.1] | 1,690 (36.4%)<br>[35.0-37.8] | 386 (40.3%)<br>[37.2-43.4] | 3,379<br>(35.5%)<br>[34.5-36.5] | 30,277<br>(28.3%)<br>[28.0-28.6] | 3,653<br>(33.2%)<br>[32.3-34.1] | 1,518<br>(27.1%)<br>[25.9-28.3] | 1,507<br>(36.7%)<br>[35.2-38.2] |
| Dispensed during pregnancy | 16,761<br>(68.1%)<br>[67.5-68.7] | 1,426 (64.5%)<br>[62.4-66.4] | 579 (70.9%)<br>[67.7-73.9] | 20,691<br>(63.0%)<br>[62.5-63.5] | 3,098 (66.8%)<br>[65.4-68.1] | 658 (68.6%)<br>[65.6-71.5] | 6,290<br>(66.1%)<br>[65.1-67.0] | 59,747<br>(55.9%)<br>[55.6-56.2] | 6,637<br>(60.3%)<br>[59.4-61.2] | 2,978<br>(53.2%)<br>[51.9-54.5] | 2,582<br>(62.9%)<br>[61.4-64.4] |
| C: Cardiovascular system |  |  |  |  |  |  |  |  |  |  |  |
| Dispensed in the year prior to pregnancy | 1,206 (4.9%)<br>[4.6-5.2] | 154 (7.0%)<br>[6.0-8.1] | 39 (4.8%) [3.5-6.5] | 1,779 (5.4%)<br>[5.2-5.7] | 269 (5.8%) [5.2-6.5] | 46 (4.8%) [3.6-6.3] | 489 (5.1%)<br>[4.7-5.6] | 5,817 (5.4%)<br>[5.3-5.6] | 680 (6.2%)<br>[5.7-6.6] | 374 (6.7%)<br>[6.1-7.4] | 237 (5.8%)<br>[5.1-6.5] |
| Dispensed during pregnancy | 1,977 (8.0%)<br>[7.7-8.4] | 190 (8.6%)<br>[7.5-9.8] | 71 (8.7%) [6.9-10.3] | 2,618 (8.0%)<br>[7.7-8.3] | 410 (8.8%) [8.1-9.7] | 70 (7.3%) [5.8-9.1] | 718 (7.5%)<br>[7.0-8.1] | 7,863 (7.4%)<br>[7.2-7.5] | 933 (8.5%)<br>[8.0-9.0] | 438 (7.8%)<br>[7.1-8.6] | 345 (8.4%)<br>[7.6-9.3] |
| D: Dermatologicals |  |  |  |  |  |  |  |  |  |  |  |
| Dispensed in the year prior to pregnancy | 9,548<br>(38.8%)<br>[38.2-39.4] | 895 (40.5%)<br>[38.4-42.5] | 319 (39.0%)<br>[35.8-42.4] | 13,354<br>(40.7%)<br>[40.1-41.2] | 1,973 (42.5%)<br>[41.1-43.9] | 368 (38.4%)<br>[35.3-41.5] | 3,404<br>(35.8%)<br>[34.8-36.7] | 43,933<br>(41.1%)<br>[40.8-41.4] | 4,489<br>(40.8%)<br>[39.9-41.7] | 2,421<br>(43.2%)<br>[41.9-44.5] | 1,726<br>(42.0%)<br>[40.5-43.6] |
| Dispensed during pregnancy | 8,559<br>(34.8%)<br>[34.2-35.4] | 798 (36.1%)<br>[34.1-38.1] | 314 (38.4%)<br>[35.2-41.8] | 12,064<br>(36.7%)<br>[36.2-37.3] | 1,855 (40.0%)<br>[38.6-41.4] | 337 (35.1%)<br>[32.2-38.2] | 3,087<br>(32.4%)<br>[31.5-33.4] | 39,843<br>(37.3%)<br>[37.0-37.5] | 4,107<br>(37.3%)<br>[36.4-38.2] | 2,192<br>(39.1%)<br>[37.9-40.4] | 1,557<br>(37.9%)<br>[36.5-39.4] |
| G: Genito urinary system and sex hormones |  |  |  |  |  |  |  |  |  |  |  |
| Dispensed in the year prior to pregnancy | 11,626<br>(47.2%)<br>[46.6-47.9] | 1,020 (46.1%)<br>[44.0-48.2] | 394 (48.2%)<br>[44.8-51.7] | 16,915<br>(51.5%)<br>[51.0-52.1] | 2,478 (53.4%)<br>[52.0-54.8] | 458 (47.8%)<br>[44.6-50.9] | 5,034<br>(52.9%)<br>[51.9-53.9] | 57,581<br>(53.8%)<br>[53.5-54.1] | 6,334<br>(57.6%)<br>[56.6-58.5] | 3,277<br>(58.5%)<br>[57.2-59.8] | 2,138<br>(52.1%)<br>[50.6-53.6] |
| Dispensed during pregnancy | 9,243<br>(37.6%)<br>[37.0-38.2] | 788 (35.6%)<br>[33.7-37.6] | 340 (41.6%)<br>[38.3-45.0] | 12,741<br>(38.8%)<br>[38.3-39.3] | 1,887 (40.7%)<br>[39.3-42.1] | 351 (36.6%)<br>[33.6-39.7] | 3,569<br>(37.5%)<br>[36.5-38.5] | 43,582<br>(40.8%)<br>[40.5-41.0] | 4,606<br>(41.9%)<br>[40.9-42.8] | 2,366<br>(42.2%)<br>[40.9-43.5] | 1,579<br>(38.5%)<br>[37.0-40.0] |
| H: Systemic hormonal preparations, excluding sex hormones and insulins |  |  |  |  |  |  |  |  |  |  |  |
| Dispensed in the year prior to pregnancy | 7,070<br>(28.7%)<br>[28.2-29.3] | 470 (21.2%)<br>[19.6-23.0] | 213 (26.1%)<br>[23.2-29.2] | 9,383<br>(28.6%)<br>[28.1-29.1] | 1,350 (29.1%)<br>[27.8-30.4] | 255 (26.6%)<br>[23.9-29.5] | 2,473<br>(26.0%)<br>[25.1-26.9] | 27,885<br>(26.1%)<br>[25.8-26.3] | 3,118<br>(28.3%)<br>[27.5-29.2] | 1,487<br>(26.5%)<br>[25.4-27.7] | 1,136<br>(27.7%)<br>[26.3-29.1] |
| Dispensed during pregnancy | 3,626<br>(14.7%)<br>[14.3-15.2] | 246<br>(11.1%)<br>[9.9-12.5] | 111<br>(13.6%)<br>[11.4-16.1] | 4,532<br>(13.8%)<br>[13.4-14.2] | 687<br>(14.8%)<br>[13.8-15.9] | 153<br>(16.0%)<br>[13.8-18.4] | 1,336<br>(14.0%)<br>[13.3-14.7] | 13,324<br>(12.5%)<br>[12.3-12.7] | 1,609<br>(14.6%)<br>[14.0-15.3] | 709<br>(12.7%)<br>[11.8-13.6] | 601<br>(14.6%)<br>[13.6-15.8] |
| **J: Antiinfectives for systemic use** |  |  |  |  |  |  |  |  |  |  |  |
| Dispensed in the year prior to pregnancy | 14,720<br>(59.8%)<br>[59.2-60.4] | 1,224 (55.3%)<br>[53.3-57.4] | 486 (59.5%)<br>[56.1-62.8] | 20,096<br>(57.3%)<br>[56.8-57.9] | 2,890 (62.3%)<br>[60.9-63.7] | 560 (58.4%)<br>[55.2-61.5] | 5,284<br>(55.5%)<br>[54.5-56.5] | 64,476<br>(60.3%)<br>[60.0-60.6] | 6,867<br>(62.4%)<br>[61.5-63.3] | 3,544<br>(63.3%)<br>[62.0-64.5] | 2,425<br>(59.1%)<br>[57.6-60.6] |
| Dispensed during pregnancy | 10,886<br>(44.2%)<br>[43.6-44.8] | 949 (42.9%)<br>[40.9-45.0] | 382 (46.8%)<br>43.4-50.2] | 14,977<br>(45.6%)<br>[45.1-46.2] | 2,161 (46.6%)<br>[45.1-48.0] | 393 (41.0%)<br>[37.9-44.1] | 4,149<br>(43.6%)<br>[42.6-44.6] | 47,728<br>(44.6%)<br>[44.3-44.9] | 5,066<br>(46.0%)<br>[45.1-47.0] | 2,571<br>(45.9%)<br>[44.6-47.2] | 1,938<br>(47.2%)<br>[45.7-48.7] |
| **L: Antineoplastic and immunomodulating agents** |  |  |  |  |  |  |  |  |  |  |  |
| Dispensed in the year prior to pregnancy | 764 (3.1%) [2.9-3.3] | 49 (2.2%) [1.7-2.9] | 22 (2.7%) [1.8-4.0] | 836 (2.5%) [2.4-2.7] | 110 (2.4%) [2.0-2.8] | 31 (3.2%) [2.3-4.6] | 267 (2.8%) [2.5-3.2] | 1,978 (1.8%) [1.8-1.9] | 214 (1.9%) [1.7-2.2] | 90 (1.6%) [1.3-2.0] | 101 (2.5%) [2.0-3.0] |
| Dispensed during pregnancy | 183 (0.7%) [0.6-0.9] | 11 (0.5%) [0.3-0.9] | < 10 | 218 (0.7%) [0.6-0.8] | 28 (0.6%) [0.4-0.9] | < 10 | 61 (0.6%) [0.5-0.8] | 501 (0.5%) [0.4-0.5] | 54 (0.5%) [0.4-0.6] | 25 (0.4%) [0.3-0.7] | 20 (0.5%) [0.3-0.8] |
| **M: Musculo-skeletal system** |  |  |  |  |  |  |  |  |  |  |  |
| Dispensed in the year prior to pregnancy | 10,874<br>(44.2%)<br>[43.6-44.8] | 746 (33.7%)<br>[31.8-35.7] | 354 (43.3%)<br>[40.0-46.8] | 16,488<br>(50.2%)<br>[49.7-50.8] | 2,375 (51.2%)<br>[49.7-52.6] | 394 (41.1%)<br>[38.0-44.2] | 3,989<br>(41.9%)<br>[40.9-42.9] | 56,878<br>(53.2%)<br>[52.9-53.5] | 6,201<br>(56.4%)<br>[55.4-57.3] | 3,358<br>(59.9%)<br>[58.7-61.2] | 2,015<br>(49.1%)<br>[47.6-50.6] |
| Dispensed during pregnancy | 2,556<br>(10.4%)<br>[10.0-10.8] | 231 (10.4%)<br>[9.2-11.8] | 84 (10.3%)<br>[8.4-12.6] | 4,310<br>(13.1%)<br>[12.8-13.5] | 593 (12.8%)<br>[11.9-13.8] | 81 (8.4%) [6.8-10.4] | 898 (9.4%)<br>[8.9-10.0] | 17,453<br>(16.3%)<br>[16.1-16.5] | 1,768<br>(16.1%)<br>[15.4-16.8] | 1,012<br>(18.1%)<br>[17.1-19.1] | 521 (12.7%)<br>[11.7-13.7] |
| **N: Nervous system** |  |  |  |  |  |  |  |  |  |  |  |
| Dispensed in the year prior to pregnancy | 16,376<br>(66.5%)<br>[65.9-67.1] | 1,322 (59.8%)<br>[57.7-61.8] | 555 (67.9%)<br>[64.7-71.0] | 23,577<br>(71.8%)<br>[71.3-72.3] | 3,451 (74.4%)<br>[73.1-75.6] | 595 (62.0%)<br>[58.9-65.1] | 5,797<br>(60.9%)<br>[59.9-61.8] | 77,984<br>(72.9%)<br>[72.7-73.2] | 8,270<br>(75.2%)<br>[74.3-76.0] | 4,331<br>(77.3%)<br>[76.2-78.4] | 2,663<br>(64.9%)<br>[63.4-66.3] |
| Dispensed during pregnancy | 14,389<br>(58.5%)<br>[57.8-59.1] | 1,140 (51.5%)<br>[49.5-53.6] | 503 (61.6%)<br>[58.2-64.8] | 20,922<br>(63.7%)<br>[63.2-64.3] | 2,997 (64.6%)<br>[63.2-66.0] | 517 (53.9%)<br>[50.7-57.0] | 5,052<br>(53.1%)<br>[52.1-54.1] | 69,059<br>(64.6%)<br>[64.3-64.9] | 7,300<br>(66.3%)<br>[65.5-67.2] | 3,699<br>(66.0%)<br>[64.8-67.3] | 2,501<br>(60.9%)<br>[59.4-62.4] |
| **P: Antiparasitic products, insecticides and repellents** |  |  |  |  |  |  |  |  |  |  |  |
| Dispensed in the year prior to pregnancy | 966 (3.9%)<br>[3.7-4.2] | 115 (5.2%)<br>[4.3-6.2] | 33 (4.0%) [2.9-5.6] | 1,593 (4.9%)<br>[4.6-5.1] | 248 (5.3%) [4.7-6.0] | 45 (4.7%) [3.5-6.2] | 421 (4.4%)<br>[4.0-4.9] | 6,331 (5.9%)<br>[5.8-6.1] | 685 (6.2%)<br>[5.8-6.7] | 393 (7.0%)<br>[6.4-7.7] | 237 (5.8%)<br>[5.1-6.5] |
| Dispensed during pregnancy | 652 (2.6%)<br>[2.5-2.9] | 73 (3.3%) [2.6-4.1] | 28 (3.4%) [2.4-4.9] | 1,028 (3.1%)<br>[2.9-3.3] | 170 (3.7%) [3.2-4.2] | 25 (2.6%) [1.8-3.8] | 295 (3.1%)<br>[2.8-3.5] | 4,324 (4.0%)<br>[3.9-4.2] | 428 (3.9%)<br>[3.5-4.3] | 269 (4.8%)<br>[4.3-5.4] | 145 (3.5%)<br>[3.0-4.1] |
| R: Respiratory system |  |  |  |  |  |  |  |  |  |  |  |
| Dispensed in the year prior to pregnancy | 11,853<br>(48.2%)<br>[47.5-48.8] | 787 (35.6%)<br>[33.6-37.6] | 397 (48.6%)<br>[45.2-52.0] | 17,394<br>(53.0%)<br>[52.4-53.5] | 2,518 (54.3%)<br>[52.8-55.7] | 426 (44.4%)<br>[41.3-47.6] | 3,949<br>(41.5%)<br>[40.5-42.5] | 55,022<br>(51.4%)<br>[51.2-51.7] | 5,651<br>(51.4%)<br>[50.4-52.3] | 2,967<br>(53.0%)<br>[51.7-54.3] | 1,967<br>(47.9%)<br>[46.4-49.4] |
| Dispensed during pregnancy | 8,783<br>(35.7%)<br>[35.1-36.3] | 634 (28.7%)<br>[26.8-30.6] | 316 (38.7%)<br>[35.4-42.1] | 12,827<br>(39.1%)<br>[38.5-39.6] | 1,848 (39.8%)<br>[38.4-41.2] | 328 (34.2%)<br>[31.3-37.3] | 3,097<br>(32.5%)<br>[31.6-33.5] | 40,251<br>(37.6%)<br>[37.3-37.9] | 4,220<br>(38.3%)<br>[37.4-39.3] | 2,110<br>(37.7%)<br>[36.4-38.9] | 1,608<br>(39.2%)<br>[37.7-40.7] |
| **S: Sensory organs** |  |  |  |  |  |  |  |  |  |  |  |
| Dispensed in the year prior to pregnancy | 4,282<br>(17.4%)<br>[16.9-17.9] | 392 (17.7%)<br>[16.2-19.4] | 166 (20.3%)<br>[17.7-23.2] | 5,566<br>(17.0%)<br>[16.6-17.4] | 774 (16.7%)<br>[15.6-17.8] | 152 (15.8%)<br>[13.7-18.3] | 1,321<br>(13.9%)<br>[13.2-14.6] | 17,169<br>(16.1%)<br>[14.8-16.3] | 1,712<br>(15.6%)<br>[14.9-16.2] | 890 (15.9%)<br>[15.0-16.9] | 785 (19.1%)<br>[17.9-20.4] |
| Dispensed during pregnancy | 2,134 (8.7%)<br>[8.3-9.0] | 223 (10.1%)<br>[8.9-11.4] | 78 (9.5%) [7.7-11.8] | 2,889 (8.8%)<br>[8.5-9.1] | 375 (8.1%) [7.3-8.9] | 83 (8.7%) [7.0-10.6] | 710 (7.5%)<br>[6.9-8.0] | 8,165 (7.6%)<br>[7.5-7.8] | 858 (7.8%)<br>[7.3-8.3] | 444 (7.9%)<br>[7.2-8.7] | 381 (9.3%)<br>[8.4-10.2] |

**Footnote.** ATC, Anatomical, Therapeutic and Chemical classification system; CI: confidence interval. Expressed in n (%) and 95% confidence interval in brackets. Differences between intra- and inter-groups (across healthcare and assistance occupations and non-healthcare occupations) were considered statistically significant when there was no overlap between the 95% CIs.

Analyses by employment type did not revealed marked differences except among nurses and hospital service workers. Folic acid dispensing before and early in pregnancy was significantly higher in the public sector than in the private sector for nurses, whereas the opposite was observed for hospital service workers (**S4 Table**). The dispensing of musculoskeletal, nervous system and respiratory medications was significantly higher among nurses and healthcare assistants working in the social assistance sector than among those working in the healthcare sector **(S5 Table)**.

### Medication dispensing during pregnancy

During pregnancy, 91% to 96% of women were dispensed at least one medication. While medication dispensing slightly declined among healthcare assistants and hospital service workers compared with the pre-pregnancy period, it remained stable in other occupations. The mean number of medications dispensed was similar across occupations (mean: 8) (**Table 2**). Occupational differences in therapeutic class exposure were less pronounced during pregnancy, but patterns persisted for nervous system, musculoskeletal and respiratory medications, which remained more frequently dispensed to social workers, psychologists, healthcare assistants, and hospital service workers (**Table 3**). Conversely, medications from alimentary tract and metabolism (ATC A) and blood and blood-forming organs (ATC B) were dispensed significantly less frequently to hospital service workers (73% [95% CI 72–74] and 53% [95% CI 52–55], respectively) compared with up to 82% [95% CI 79–84] and 69% [95% CI 66–72] among rehabilitation professionals. No significant differences were observed by employment type or sector **(S4 and S5 Tables)**.

### Specific medication classes

Folic acid dispensing from before pregnancy to the first trimester of pregnancy varied by occupation (**Figure 2a, S6 Table)**, with significantly higher rates among physicians (51% [95% CI 49–53]), psychologists (50% [95% CI 47–54]) and rehabilitation professionals (53% [95% CI 50–57]) compared with healthcare assistants (39% [95% CI 37.7–39.5] and hospital service workers (30% [95% CI 29–32)). Compared with non-healthcare workers of similar socioeconomic level, most healthcare professionals had higher folic acid dispensing, except hospital service workers. Psychotropic medication (including antidepressants and benzodiazepines) dispensing before pregnancy was significantly higher among hospital service workers (18% [95% CI 17–19]), healthcare assistants (16% [95% CI 15–16]), and social workers (14% [95% CI 13–15]) than among physicians (11% [95% CI 10–13]), rehabilitation professionals (10% [95% CI 8–12]), and nurses (10% [95% CI 9–10]) (**Figure 2b, S6 Table)**. Inter-sector comparisons showed that the former group had significantly higher dispensing rates than non-healthcare workers of similar socioeconomic level, while the latter group had significantly lower rates. Analgesic dispensing before pregnancy was significantly higher among hospital service workers (73% [95% CI 72–74]) and healthcare assistants (70% [95% CI 70–71]) than among other health professionals, with rates as low as 54% for physicians (95% CI 52–56) and nurses (95% CI 53–55), and among non-healthcare workers of similar socioeconomic level (68%) (**Figure 2c, S6 Table)**. Nurses and healthcare assistants employed in the social assistance sector exhibited significantly higher dispensing rates than those employed in the healthcare sector **(S5 Table)**.

**Figure 2.** Prevalence of specific medication dispensed by occupational groups (expressed in %) (a) Prevalence was estimated for each occupational group from the year prior to pregnancy to the end of the first trimester of pregnancy. (b) and (c) Bars with solid texture represent estimates in the year prior to pregnancy, while bars with white-dot texture represent estimates during pregnancy.

Differences in exposure to specific medication classes were less pronounced during pregnancy **(S6 Table).**

## Discussion

In this large population-based study of nearly 39,000 pregnant women working in the healthcare and social assistance sector in France, we observed substantial heterogeneity in medication dispensing across occupations. Women with lower socioeconomic levels, particularly hospital service workers and healthcare assistants, had significantly higher rates of psychotropics (including antidepressants and benzodiazepines), antalgics and musculoskeletal medication dispensing. Conversely, physicians had significantly lower medication exposure, except vitamins and supplements. These differences persisted after accounting for socioeconomic level, suggesting that occupational factors may contribute to these differences.

Before pregnancy, a study found that 91% of women in France had at least one medication dispensed, with an average of 10.8 prescriptions ^8^, which is in line with our results. During pregnancy, over 90% of women in our study received at least one dispensed medication, including iron and vitamins. This is consistent with previous estimates from France ^1^, but higher than those reported in other countries such as the Netherland (79%, including iron and vitamins) ^9^, Denmark (63%, including iron and vitamins) ^10^ and the United States (65%, excluding iron and vitamins) ^11^. However, international comparisons should be interpreted with caution, as the types of reimbursed medications may vary across countries, potentially affecting prevalence estimates. Compared with results among French pregnant women ^1^, women in our study had higher dispensing rates for ATC classes A (alimentary tract and metabolism), B (blood and blood forming organs), and R (respiratory system), but lower for classes M (musculo-skeletal system) and N (nervous system), with notable variation by occupations.

Despite working in the same sector, marked differences were observed. Workers with lower socioeconomic levels had significantly higher rates of psychotropic, analgesic, and musculoskeletal medication dispensing than worker with the higher ones. This is consistent with previous findings showing a socioeconomic gradient. For instance, a study reported that low-income pregnant women were less likely to receive recommended prescriptions, such as vitamins during pregnancy, while analgesics, fetotoxic medications, and medications related to chronic diseases were more frequently dispensed in this population ^12^. Similarly, another study investigating medication consumption among French workers found that executives used less analgesics and musculoskeletal medications than intermediate, employees and manual workers ^13^. Our results extend these observations by demonstrating that such gradients persist even within the healthcare sector.

These disparities likely reflect a combination of factors, including differences in working conditions, health status, education, and health literacy. Chronic diseases are more prevalent among women in lower socioeconomic levels ^14^, and could partly explain the greater need for medications. Higher folic acid dispensing among physicians, rehabilitation professionals and nurses suggests greater awareness of prenatal care recommendations, whereas lower rates among healthcare assistants and hospital service workers point to gaps in preventive counseling, despite their proximity to the healthcare system. These findings align with studies linking low health literacy to higher rate of unplanned pregnancies, a lower use of folic acid prior or during pregnancy ^15^. Moreover, this study reported a higher rate of hospitalizations during pregnancy among women with low health literacy compared with those of adequate health literacy ^15,16^.

Occupational exposures in healthcare such as shift work, physical demands, and emotional stress may contribute to these differences beyond socioeconomic factors. Occupations involving shift work and physical demands (such as hospital service workers and assistants) showed the highest medication exposure. Previous research has shown that such working conditions are associated with increased use of psychotropic, musculoskeletal and digestive medications, as well as poorer self-reported health.^17–20^. Elevated rates of psychotropic and analgesic medication dispensing among hospital service workers, psychologists and social workers in our study likely reflect cumulative occupational stress. Specifically, in a French workforce survey, perceived difficult working conditions was associated with increased use of anxiolytic, psychotropic, musculoskeletal and digestive medications ^13^.

Medication dispensing declined during pregnancy, likely reflecting increased awareness of foetal risks and specialist pregnancy-related care. Nevertheless, elevated exposure persisted in certain occupational groups, possibly due to greater burden of pre-existing conditions or limited access to non-pharmacological alternatives.

### Strengths and limitations

This study has several strengths, including the use of a large, national cohort linking occupational information with healthcare data. The granularity of occupational information enabled nuanced comparisons across occupational groups, highlighting why examining overall averages may obscure important heterogeneity: aggregating all healthcare professionals would miss the higher medication exposure in vulnerable occupational subgroups. The linkage to the SNDS provided robust and detailed data on medication dispensing. However, several limitations should be acknowledged. First, medication data were limited to prescribed, reimbursed and dispensed out-patient prescriptions, and did not capture over-the-counter (OTC) medications or in-hospital medications. We cannot rule out a differential use of OTC medications among the healthcare occupations, due to differences in pharmacological knowledge and/or financial resources. Second, differences were assessed using non-overlapping confidence intervals, a conservative approach that may have missed small effects, although such differences would be unlikely to translate into clinically meaningful effects. Third, some healthcare occupations may influence medication dispensing patterns through specific professional roles (i.e prescriber status), while direct workplace access to medications may lead to under-recording in administrative data. Fourth, we lacked detailed information on working conditions, such as shift work or physically demanding activities, which may have contributed to health differences. Fifth, the data were collected more than ten years ago. However, the main patterns are likely to remain valid. Nevertheless, given the worsening of working conditions in healthcare sectors in recent years, our findings are unlikely to be attenuated in more recent cohorts. Finally, because our study included mainly salaried women, our results may not be generalizable to self-employed workers.

### Perspectives for future research and implications

Future studies should explore how specific work conditions (shift work, high patient load, exposure to infectious agents, and ergonomic constraints) contribute to the disparities in maternal health and pregnancy outcomes among healthcare professionals. Our findings support the need for targeted preventive strategies for healthcare and social assistance workers, particularly in high-risk occupations like hospital service workers and assistants. Interventions should address working conditions, improve access to preventive care, and tailor occupation-specific support programs to reduce intra-sector disparities.

### Conclusion

In this large French population-based study of nearly 40,000 pregnant women working in the healthcare and social assistance sector, we observed clear occupational gradients in medication dispensing during pregnancy within the healthcare and social assistance sector: women with lower socioeconomic level, such as hospital service workers and healthcare assistants, had higher exposure to psychotropics, antalgics and musculoskeletal medications and lower folic acid supplementation. These disparities persisted after accounting for socioeconomic level, underscoring the importance of occupation-specific factors, such as working conditions, educational level and health literacy, on maternal health. Further studies will examine how work-related strain impacts both the health of these women and their pregnancy outcomes.

## Declarations Ethic statement

Our study was approved by the Statistical Confidentiality Committee on November, 30 2023, and by the French National Archives on December 19, 2023. Additionally, approval was obtained from the Ethics and Scientific Committee for Research, Studies, and Evaluations in the Health Field (CESREES) on October, 19 2023, and from the French data protection authority (CNIL) on February, 23 2024 (agreement number: DR-2024-037). The study involved the secondary use of pseudonymized health and socio-economic data. In accordance with national regulations and ethical guidelines, individual informed consent was not required. An exception to the obligation of individual information were made, in compliance with Article 14 of the General Data Protection Regulation. All data were pseudonymized prior to analysis, and researchers did not have access to any directly identifiable information.

## Data availability

The data cannot be shared publicly as it contains sensitive, individual health data. Restrictions apply to the availability of these data, which were used under license for this study. Data are owned by a third-party organization and access to the data set are conditioning upon approval from the owner (Department for Research, Studies, Assessment and Statistics) and from the “Statistical Confidentiality Committee”, the “French National Archives”, the “Ethics and Scientific Committee for Research, Studies, and Evaluations in the Health Field”, and the “French data protection authority”. Data requests can be sent to.

## Authors contribution

All authors contributed to the study conception and design. Material preparation, data collection and analysis were performed by Mélanie Araujo. The first draft of the manuscript was written by Anna-Belle Beau and all authors commented on previous versions of the manuscript. All authors read and approved the final manuscript.

## Supporting information captions

**<u>S1 Methods</u>:** Description of the data source EDP-Santé

**<u>S1 Table</u>:** Pregnancy identification algorithm

**<u>S2 Table</u>:** Occupational classification and codes

**<u>S3 Table</u>:** Anatomical Therapeutic Chemical (ATC) codes used to classify medication exposure

**<u>S4 Table</u>:** Stratification by type of employment (public *versus* private): prevalence of medication dispensed in the year prior to and during pregnancy by occupational groups

**<u>S5 Table</u>:** Stratification by sector of employment (healthcare *versus* social assistance): prevalence of medication dispensed in the year prior to and during pregnancy by occupational groups

**<u>S6 Table</u>:** Prevalence of specific medication dispensed in the year prior to and during pregnancy by occupational groups

