## Supplementary Materials for "Drug utilization across occupations among pregnant women working in the healthcare and social assistance sector in France: a nationwide population-based study"

**S 1 Methods: Description of the data source EDP-Santé**

The study utilized data from the EDP-Santé 2017, a French administrative database linking two major data sources: the permanent demographic sample (*Echantillon Démographique de la Population [EDP]*) from the French National Institute for Statistics and Economic Studies (*Institut National de la Statistique et des Études Économiques [INSEE]*) and the National Health Data System (*Système National des Données de Santé [SNDS]*) made available by the French national healthcare insurance system (*Caisse Nationale de l'Assurance Maladie [CNAM]*). Briefly, the EDP contains socioeconomic data, mainly derived from tax declaration, employer declaration (private and public sectors) and annual census records. Self-employed individuals can only be identified from the annual census records. The SNDS contains comprehensive information on healthcare utilization, including both outpatient (medications, medical procedures from the French national health insurance database, *Datamart de Consomation Inter-Régime [DCIR]*) and inpatient (diagnosis codes, expensive medications and medical procedures from the French hospital discharge database, *Programme de Médicalisation des Systèmes d'Information* [PMSI]) data. Diseases are coded according to the International Classification of Diseases, 10$^{th}$ Revision (ICD-10), and medical procedures are coded according to the French medical classification for clinical procedures (CCAM). The linkage between these two data sources was made possible through a unique national identification number, assigned at birth as part of the national membership registry and commonly referred to as the "social security number". EDP-Santé constitutes a representative sample of the French population, covering approximately 4% of individuals.

**S1 Table: Pregnancy identification algorithm**

Pregnancy episodes were identified using the SNDS through hospital discharge diagnoses coded according to the ICD-10 and medical procedures coded according to the CCAM from public and private establishments. Outpatient medical abortions were identified from the DCIR database and all other pregnancy types from the PMSI-MCO (Medicine, Surgery, Obstetrics and Odontology) database. Diagnoses and procedures are presented in the following table. The start of pregnancy was estimated based on the pregnancy end date (exact admission, discharge and medical procedure dates recorded in the PMSI MCO database) and the gestational age or number of days since the last menstrual period (LMP), as recorded in the PMSI-MCO database.

| Pregnancy outcomes | Identification codes | Median gestational age (in weeks)<br>*(source: 2012 Abortion Bulletins - INED)* |
|---|---|---|
| Stillbirths | Associated diagnoses Z37.10. Z37.30. Z37.40. Z37.60. Z37.70 | 30 |
| Elective abortions | Principal diagnoses O04. O05. O06. O07<br>AND procedure indicative of inpatient abortion. CCAM codes: JNJD001. JNJD002. JNJP001<br>AND Associated diagnosis Z640<br><br>OR<br><br>Procedure indicative of outpatient medical abortion. CCAM codes: 1981. 2411. 2414. 2419. 2420. 2422. 2423. 2425. 2426. 3329 | Procedure indicative of inpatient abortion: 7,<br>Procedure indicative of inpatient abortion in the first trimester of pregnancy: 9,<br>Procedure indicative of inpatient abortion in the second trimester of pregnancy: 13<br><br>Procedure indicative of outpatient medical abortion: 6 |
| Therapeutic abortions<br>< 22 weeks after the LMP | Principal diagnoses O04. O05. O06. O07<br>AND procedure indicative of inpatient abortion. CCAM codes: JNJD001. JNJD002. JNJP001<br>WITHOUT Associated diagnosis Z640 | Medical therapeutic abortion: 14,<br>Therapeutic abortion in the first trimester of pregnancy: 10,<br>Therapeutic abortion in the second trimester of pregnancy: 17 |
| Therapeutic abortions<br>≥ 22 weeks after the LMP | Associated diagnoses Z37.11. Z37.31. Z37.41. Z37.61. Z37.71 | 26 |
| Spontaneous abortions | Principal diagnosis O03 | 9 |
| Other abortions | Principal diagnoses O04. O05. O06. O07 | 9 |

| | | |
|---|---|---|
| | WITHOUT CCAM codes: JNJD001. JNJD002. JNJP001 | |
| Ectopic pregnancies | Principal diagnosis O00<br>OR procedure indicative of inpatient abortion. CCAM codes: JJFA001. JJFC001. JJJA002. JJJC002. JJLJ001. JJPA001. JJPC001. JQGA001 | 6 |
| Hydatidiform mole | Principal diagnosis O01 | 9 |
| Other abnormal products of conception | Principal diagnosis O02 | 9 |
| Live births | Principal diagnoses O80. O81. O82. O83. O84<br>OR Associated diagnoses Z37. Z3900<br>OR CCAM codes JQGD001 to 005. JQGD007. JQGD008. JQGD010. JQGD012. JQGD013. JQGA002-005<br>WITHOUT Stillbirths OR Therapeutic abortions ≥ 22 weeks of amenorrhea | 39 |

**Footnote:** If not otherwise specified, the codes are ICD-10 codes, corresponding to hospital discharge diagnoses. CCAM, French medical classification of clinical procedures; ICD-10, International Classification of Disease, 10th revision.

**S2 Table : Occupational classification and codes**

Based on the 2003 French Classification of Occupations and Socioprofessional Categories, level 4 (CSP 2003).

| **Socioeconomic level** | **Occupational category** | **Occupations** | **CSP 2003 codes** |
|---|---|---|---|
| **Used for exclusion** | | | |
| Farmers | | | Starting by 1 |
| Artisans/traders, company managers | | | Starting by 2 |
| Laborers | | | Starting by 6 |
| **Used for inclusion and classification** | | | |
| Executive | physicians | physicians, dental surgeons and midwives | 311A, 311B, 311C, 344A, 344B, 344C and 431E |
| | psychologists | clinical and occupational psychologists | 311D and 343A |
| Intermediate | nurses | general care nurses, specialized nurses, operating room nurses, etc. | 431A, 431B, 431C, 431D, 431F and 431G |
| | social workers | social workers, family counselors, etc. | 434 |
| | rehabilitation professionals | physiotherapists, occupational therapists, psychomotor therapists | 432 |
| Employees | healthcare assistants | nursing, dental or medical or psychologist assistants, and, childcare auxiliaries | 526A, 526B, 526C and 526D |
| | hospital service workers | hospital cleaning, logistics | 525 |
| NA | others healthcare professionals, not classified elsewhere | pharmacists, medical laboratory technicians, dietitians, etc. | 311F, 344D, 433A, 433B, 433C, 433D and 526E |

**S3 Table : Anatomical Therapeutic Chemical (ATC) codes used to classify medication exposure**

| Medication class | ATC codes included |
|---|---|
| Folic acid | B03AD03, B03BB01, B05XC |
| Psychotropic medications | N03, N05A, N05BA, N05CD, N05CF, N06A |
| Benzodiazepines | N05BA, N05CD, N05CF |
| Antidepressants | N06A |
| Analgesics (excluding antimigraine drugs) | N02, excluding N02C |
| Musculoskeletal medications | M01, M02, M03, M09AX01, M09AX02 |
| NSAIDs | M01A (except M01AX26), M01B, M02A, N02BA01/10/15/51/65/71 |

**S4 Table : Stratification by type of employment (public versus private): prevalence of medication dispensed in the year prior to and during pregnancy by occupational groups.**

| | Executives | | | | Intermediate occupations | | | | | | Employees | | | |
|---|---|---|---|---|---|---|---|---|---|---|---|---|---|---|
| | PhysiciansN=2,212 | | Psychologists N=817 | | Nurses N=9,521 | | Social workers N=4,640 | | Rehabilitation professionals N=959 | | Healthcare Assistants N=11,004 | | Hospital service workers N=5,602 | |
| | ***Public*** *N=1,735* | ***Private*** *N=399* | ***Public*** *N=389* | ***Private*** *N=393* | ***Public*** *N=6,130* | ***Private*** *N=3,222* | ***Public*** *N=1,556* | ***Private*** *N=2,947* | ***Public*** *N=324* | ***Private*** *N=487* | ***Public*** *N=5,086* | ***Private*** *N=5,748* | ***Public*** *N=2,599* | ***Private*** *N=2,841* |
| **DISPENSED MEDICATIONS** | | | | | | | | | | | | | | |
| **Medication dispensed in the year prior to pregnancy** | | | | | | | | | | | | | | |
| **At least one medication** [a] | 1,583 (91.2%) [89.8-92.5] | 361 (90.5%) [87.2-93.0] | 369 (94.9%) [92.2-96.7] | 374 (95.2%) [92.6-96.9] | 5,714 (93.2%) [92.6-93.8] | 3,007 (93.3%) [92.4-94.1] | 1,487 (95.6%) [94.4-96.5] | 2,811 (95.4%) [94.6-96.1] | 310 (95.7%) [92.9-97.4] | 449 (92.2%) [89.5-94.3] | 4,847 (95.3%) [94.7-95.9] | 5,474 (95.2%) [94.7-95.8] | 2,445 (94.1%) [93.1-94.9] | 2,644 (93.1%) [92.1-93.9] |
| **Number of different medications** [b] | 8.0±6.6 [7.7-8.3] | 8.8±6.8 [8.2-9.5] | 9.4±6.9 [8.8-10.1] | 9.6±6.5 [8.9-10.2] | 8.6±6.6 [8.4-8.7] | 8.8±6.7 [8.6-9.1] | 10.5±7.3 [10.2-10.9] | 11.0±7.8 [10.7-11.2] | 9.5±7.4 [8.7-10.3] | 9.0±7.1 [8.4-9.7] | 11.1±8.0 [10.8-11.3] | 11.5±8.4 [11.2-11.7] | 12.2±9.2 [11.8-12.5] | 12.1±9.2 [11.8-12.5] |
| **A: Alimentary tract and metabolism** [a] | 854 (49.2%) [46.9-51.6] | 217 (54.4%) [49.5-59.2] | 221 (56.8%) [51.8-61.6] | 236 (60.1%) [55.1-64.8] | 3,004 (49.0%) [47.8-50.3] | 1,615 (50.1%) [48.4-51.8] | 931 (59.8%) [57.4-62.2] | 1,744 (59.2%) [57.4-60.9] | 176 (54.3%) [48.9-59.7] | 251 (51.5%) [47.1-55.9] | 2,995 (58.9%) [57.5-60.2] | 3,476 (60.5%) [59.2-61.7] | 1,604 (61.7%) [59.8-63.6] | 1,748 (61.5%) [59.7-63.3] |
| **B: Blood and blood forming organs** [a] | 715 (41.2%) [38.9-43.5] | 158 (39.6%) [34.9-44.5] | 149 (38.3%) [33.6-43.2] | 157 (39.9%) [35.2-44.9] | 2,254 (36.8%) [35.6-38.0] | 1,073 (33.3%) [31.7-34.9] | 610 (39.2%) [36.8-41.7] | 1,047 (35.5%) [33.8-37.3] | 139 (42.9%) [37.6-48.3] | 190 (39.0%) [34.8-43.4] | 1,725 (33.9%) [32.6-35.2] | 1,885 (32.8%) [31.6-34.0] | 697 (26.8%) [25.1-28.6] | 778 (27.4%) [25.8-29.1] |
| **M: Musculo-skeletal system** [a] | 569 (32.8%) [30.6-35.0] | 149 (37.3%) [32.7-42.2] | 165 (42.4%) [37.6-47.4] | 171 (43.5%) [38.7-48.5] | 2,516 (41.0%) [39.8-42.3] | 1,398 (43.4%) [41.7-45.1] | 772 (49.6%) [47.1-52.1] | 1,538 (52.2%) [50.4-54.0] | 123 (38.0%) [32.8-43.4] | 209 (42.9%) [38.6-47.4] | 2,839 (55.8%) [54.5-57.2] | 3,271 (56.9%) [55.6-58.2] | 1,573 (60.5%) [58.6-62.4] | 1,706 (60.0%) [58.2-61.8] |
| **N: Nervous system** [a] | 1,020 (58.8%) [56.5-61.1] | 250 (62.7%) [57.8-67.3] | 259 (66.6%) [61.8-71.1] | 270 (68.7%) [64.0-73.1] | 3,710 (60.5%) [59.3-61.7] | 1,981 (61.5%) [59.8-63.1] | 1,133 (72.8%) [70.6-75.0] | 2,222 (75.4%) [73.8-76.9] | 211 (65.1%) [59.8-70.1] | 303 (62.2%) [57.8-66.4] | 3,793 (74.6%) [73.4-75.8] | 4,358 (75.8%) [74.7-76.9] | 2,026 (78.0%) [76.3-79.5] | 2,183 (76.8%) [75.3-78.4] |
| **R: Respiratory system** [a] | 600 (34.6%) [32.4-36.9] | 155 (38.8%) [34.2-43.7] | 186 (47.8%) [42.9-52.8] | 190 (48.3%) [43.4-53.3] | 2,510 (40.9%) [39.7-42.2] | 1,368 (42.5%) [40.8-44.2] | 829 (53.3%) [50.8-55.7] | 1,610 (54.6%) [52.8-56.4] | 152 (46.9%) [41.5-52.4] | 214 (43.9%) [39.6-48.4] | 2,588 (50.9%) [49.5-52.3] | 2,990 (52.0%) [50.7-53.3] | 1,390 (53.5%) [51.6-55.4] | 1,486 (52.3%) [50.5-54.1] |
| **Folic acid** [a] (Dispensed in the year prior to pregnancy | 882 (50.8%) | 198 (49.6%) | 211 (54.2%) | 184 (46.8%) | 2,850 (46.5%) | 1,358 (42.1%) | 772 (49.6%) | 1,311 (44.5%) | 178 (54.9%) | 257 (52.8%) | 2,052 (40.3%) | 2,149 (37.4%) | 731 (28.1%) | 916 (32.2%) |

| | | | | | | | | | | | | | | |
|---|---|---|---|---|---|---|---|---|---|---|---|---|---|---|
| to 1st trimester of pregnancy) | [48.5-53.2] | [44.7-54.5] | [49.3-59.1] | [41.9-51.8] | [45.2-47.7] | [40.5-43.9] | [47.1-52.1] | [42.7-46.3] | [49.5-60.3] | [48.3-57.2] | [39.0-41.7] | [36.1-38.6] | [26.4-29.9] | [30.5-34.0] |
| **Psychotropic medications** [a] | 185 (10.7%) [9.3-12.2] | 47 (11.8%) [9.0-15.3] | 41 (10.5%) [7.9-14.0] | 58 (14.8%) [11.6-18.6] | 593 (9.7%) [9.0-10.4] | 315 (9.8%) [8.8-10.9] | 189 (12.1%) [10.6-13.9] | 445 (15.1%) [13.9-16.4] | 30 (9.3%) [6.6-12.9] | 52 (10.7%) [8.2-13.7] | 781 (15.4%) [14.4-16.4] | 921 (16.0%) [15.1-17.0] | 502 (19.3%) [17.8-20.9] | 498 (17.5%) [16.2-19.0] |
| **Benzodiazepines** [a] | 146 (8.4%) [7.2-9.8] | 33 (8.3%) [5.9-11.4] | 30 (7.7%) [5.5-10.8] | 49 (12.5%) [9.6-16.1] | 437 (7.1%) [6.5-7.8] | 250 (7.8%) [6.9-8.7] | 135 (8.7%) [7.4-10.2] | 355 (12.0%) [10.9-13.3] | 20 (6.2%) [4.0-9.3] | 35 (7.2%) [5.2-9.8] | 611 (12.0%) [11.1-12.9] | 739 (12.9%) [12.0-13.7] | 394 (15.2%) [13.8-16.6] | 378 (13.3%) [12.1-14.6] |
| **Antidepressants** [a] | 74 (4.3%) [3.4-5.3] | 22 (5.5%) [3.7-8.2] | 22 (5.7%) [3.8-8.4] | 19 (4.8%) [3.1-7.4] | 269 (4.4%) [3.9-4.9] | 131 (4.1%) [3.4-4.8] | 97 (6.2%) [5.1-7.5] | 205 (7.0%) [6.1-7.9] | 17 (5.2%) [3.3-8.2] | 22 (4.5%) [3.0-6.7] | 358 (7.0%) [6.4-7.8] | 427 (7.4%) [6.8-8.1] | 246 (9.5%) [8.4-10.7] | 232 (8.2%) [7.2-9.2] |
| **Analgesics** [a] | 907 (52.3%) [49.9-54.6] | 230 (57.6%) [52.7-62.4] | 234 (60.2%) [55.2-64.9] | 252 (64.1%) [59.3-68.7] | 3,321 (54.2%) [52.9-55.4] | 1,771 (55.0%) [53.2-56.7] | 1,046 (67.2%) [64.9-69.5] | 2,067 (70.1%) [68.5-71.8] | 198 (61.1%) [55.7-66.3] | 280 (57.5%) [53.1-61.8] | 3,547 (69.7%) [68.5-71.0] | 4,082 (71.0%) [69.8-72.2] | 1,913 (73.6%) [71.9-75.3] | 2,076 (73.1%) [71.4-74.7] |
| **Musculoskeletal medications** [a] | 566 (32.6%) [30.5-34.9] | 146 (36.6%) [32.0-41.4] | 164 (42.2%) [37.4-47.1] | 171 (43.5%) [38.7-48.5] | 2,508 (40.9%) [39.7-42.1] | 1,392 (43.2%) [41.5-44.9] | 770 (49.5%) [47.0-52.0] | 1,533 (52.0%) [50.2-53.8] | 123 (38.0%) [32.8-43.4] | 209 (42.9%) [38.6-47.4] | 2,831 (55.7%) [54.3-57.0] | 3,265 (56.8%) [55.5-58.1] | 1,571 (60.4%) [58.6-62.3] | 1,704 (60.0%) [58.2-61.8] |
| **NSAIDs** [a] | 573 (33.0%) [30.9-35.3] | 150 (37.6%) [33.0-42.4] | 164 (42.2%) [37.4-47.1] | 171 (43.5%) [38.7-48.5] | 2,535 (41.4%) [40.1-42.6] | 1,404 (43.6%) [41.9-45.3] | 770 (49.5%) [47.0-52.0] | 1,537 (52.2%) [50.3-54.0] | 125 (38.6%) [33.4-44.0] | 206 (42.3%) [38.0-46.7] | 2,838 (55.8%) [54.4-57.2] | 3,271 (56.9%) [55.6-58.2] | 1,558 (59.9%) [58.0-61.8] | 1,692 (59.6%) [57.7-61.3] |
| **Medication dispensed during pregnancy** | | | | | | | | | | | | | | |
| **At least one medication** [a] | 1,625 (93.7%) [92.4-94.7] | 376 (94.2%) [91.5-96.1] | 376 (96.7%) [94.4-98.0] | 382 (97.2%) [95.1-98.4] | 5,796 (94.6%) [94.0-95.1] | 3,026 (93.9%) [93.0-94.7] | 1,493 (96.0%) [94.9-96.8] | 2,808 (95.3%) [94.5-96.0] | 315 (97.2%) [94.8-98.5] | 465 (95.5%) [93.3-97.0] | 4,789 (94.2%) [93.5-94.8] | 5,395 (93.9%) [93.2-94.4] | 2,365 (91.0%) [89.8-92.0] | 2,606 (91.7%) [90.7-92.7] |
| **Number of different medications** [b] | 7.1±5.0 [6.8-7.3] | 7.5±5.7 [6.9-8.1] | 8.4±5.4 [7.8-8.9] | 8.3±5.5 [7.7-8.8] | 7.2±4.9 [7.1-7.3] | 7.3±5.0 [7.1-7.5] | 8.3±5.4 [8.0-8.5] | 8.4±5.5 [8.2-8.6] | 7.7±5.0 [7.2-8.3] | 7.5±4.8 [7.1-8.0] | 8.2±5.6 [8.0-8.3] | 8.3±5.9 [8.1-8.4] | 8.0±6.2 [7.7-8.2] | 8.3±6.1 [8.0-8.5] |
| **A: Alimentary tract and metabolism** [a] | 1,375 (79.3%) [77.3-81.1] | 308 (77.2%) [72.8-81.0] | 329 (84.6%) [80.7-87.8] | 326 (83.0%) [78.9-86.3] | 4,848 (79.1%) [78.1-80.1] | 2,537 (78.7%) [77.3-80.1] | 1,261 (81.0%) [79.0-82.9] | 2,350 (79.7%) [78.3-81.2] | 271 (83.6%) [79.2-87.3] | 402 (82.5%) [78.9-85.7] | 4,004 (78.7%) [77.6-79.8] | 4,402 (76.6%) [75.5-77.7] | 1,873 (72.1%) [70.3-73.8] | 2,102 (74.0%) [72.3-75.6] |
| **B: Blood and blood forming organs** [a] | 1,132 (65.2%) [63.0-67.4] | 248 (62.2%) [57.3-66.8] | 286 (73.5%) [68.9-77.7] | 275 (70.0%) [65.3-74.3] | 4,117 (67.2%) [66.0-68.3] | 2,074 (64.4%) [62.7-66.0] | 1,069 (68.7%) [66.4-71.0] | 1,944 (66.0%) [64.2-67.7] | 226 (69.8%) [64.5-74.5] | 342 (70.2%) [66.0-74.1] | 3,192 (62.8%) [61.4-64.1] | 3,343 (58.2%) [56.9-59.4] | 1,336 (51.4%) [49.5-53.3] | 1,551 (54.6%) [52.8-56.4] |

| | | | | | | | | | | | | | | |
|---|---|---|---|---|---|---|---|---|---|---|---|---|---|---|
| **M: Musculo-skeletal system** [a] | 185 (10.7%) [9.3-12.2] | 40 (10.0%) [7.4-13.4] | 39 (10.0%) [7.4-13.4] | 40 (10.2%) [7.6-13.6] | 559 (9.1%) [8.4-9.9] | 315 (9.8%) [8.8-10.9] | 195 (12.5%) [11.0-14.3] | 376 (12.8%) [11.6-14.0] | 23 (7.1%) [4.8-10.4] | 46 (9.4%) [7.2-12.4] | 752 (14.8%) [13.8-15.8] | 995 (17.3%) [16.4-18.3] | 451 (17.4%) [15.9-18.9] | 535 (18.8%) [17.4-20.3] |
| **N: Nervous system** [a] | 884 (51.0%) [48.6-53.3] | 212 (53.1%) [48.2-58.0] | 239 (61.4%) [56.5-66.1] | 243 (61.8%) [56.9-66.5] | 3,272 (53.4%) [52.1-54.6] | 1,691 (52.5%) [50.8-54.2] | 963 (61.9%) [59.4-64.3] | 1,948 (66.1%) [64.4-67.8] | 193 (59.6%) [54.1-64.8] | 252 (51.7%) [47.3-56.2] | 3,360 (66.1%) [64.8-67.4] | 3,831 (66.6%) [65.4-67.9] | 1,687 (64.9%) [63.1-66.7] | 1,904 (67.0%) [65.3-68.7] |
| **R: Respiratory system** [a] | 481 (27.7%) [25.7-29.9] | 128 (32.1%) [27.7-36.8] | 145 (37.3%) [32.6-42.2] | 156 (39.7%) [35.0-44.6] | 1,968 (32.1%) [30.9-33.3] | 1,074 (33.3%) [31.7-35.0] | 632 (40.6%) [38.2-43.1] | 1,166 (39.6%) [37.8-41.3] | 122 (37.7%) [32.6-43.0] | 163 (33.5%) [29.4-37.8] | 1,953 (38.4%) [37.1-39.7] | 2,201 (38.3%) [37.0-39.6] | 967 (37.2%) [35.4-39.1] | 1, 072 (37.7%) [36.0-39.5] |
| **Psychotropic medications** [a] | 79 (4.6%) [3.7-5.6] | 22 (5.5%) [3.7-8.2] | 19 (4.9%) [3.1-7.5] | 19 (4.8%) [3.1-7.4] | 221 (3.6%) [3.2-4.1] | 139 (4.3%) [3.7-5.1] | 73 (4.7%) [3.7-5.9] | 174 (5.9%) [5.1-6.8] | 14 (4.3%) [2.6-7.1] | 15 (3.1%) [1.9-5.0] | 316 (6.2%) [5.6-6.9] | 353 (6.1%) [5.5-6.8] | 179 (6.9%) [6.0-7.9] | 178 (6.3%) [5.4-7.2] |
| **Benzodiazepines** [a] | 58 (3.3%) [2.6-4.3] | 15 (3.8%) [2.3-6.1] | 11 (2.8%) [1.6-5.0] | 14 (3.6%) [2.1-5.9] | 123 (2.0%) [1.7-2.4] | 90 (2.8%) [2.3-3.4] | 43 (2.8%) [2.1-3.7] | 108 (3.7%) [3.0-4.4] | < 10 | < 10 | 205 (4.0%) [3.5-4.6] | 223 (3.9%) [3.4-4.4] | 129 (5.0%) [4.2-5.9] | 124 (4.4%) [3.7-5.2] |
| **Antidepressants** [a] | NA | NA | NA | NA | 93 (1.5%) [1.2-1.9] | 47 (1.5%) [1.1-1.9] | 32 (2.1%) [1.5-2.9] | 88 (3.0%) [2.4-3.7] | < 10 | < 10 | 133 (2.6%) [2.2-3.1] | 141 (2.5%) [2.1-2.9] | 77 (3.0%) [2.4-3.7] | 68 (2.4%) [1.9-3.0] |
| **Analgesics** [a] | 811 (46.7%) [44.4-49.1] | 197 (49.4%) [44.5-54.3] | 231 (59.4%) [54.4-64.1] | 229 (58.3%) [53.3-63.0] | 3,078 (50.2%) [49.0-51.5] | 1,595 (49.5%) [47.8-51.2] | 927 (59.6%) [57.1-62.0] | 1,837 (62.3%) [60.6-64.1] | 188 (58.0%) [52.6-63.3] | 240 (49.3%) [44.9-53.7] | 3,191 (62.7%) [61.4-64.1] | 3,667 (63.8%) [62.5-65.0] | 1,609 (61.9%) [60.0-63.8] | 1,833 (64.5%) [62.7-66.3] |
| **Musculoskeletal medications** [a] | 184 (10.6%) [9.2-12.1] | 37 (9.3%) [6.8-12.5] | 39 (10.0%) [7.4-13.4] | 38 (9.7%) [7.1-13.0] | 553 (9.0%) [8.3-9.8] | 310 (9.6%) [8.7-10.7] | 192 (12.3%) [10.8-14.1] | 370 (12.6%) [11.4-13.8] | 22 (6.8%) [4.5-10.1] | 46 (9.4%) [7.2-12.4] | 743 (14.6%) [13.7-15.6] | 987 (17.2%) [16.2-18.2] | 449 (17.3%) [15.9-18.8] | 534 (18.8%) [17.4-20.3] |
| **NSAIDs** [a] | 247 (14.2%) [12.7-16.0] | 66 (16.5%) [13.2-20.5] | 50 (12.9%) [9.9-16.5] | 47 (12.0%) [9.1-15.5] | 705 (11.5%) [10.7-12.3] | 375 (11.6%) [10.6-12.8] | 215 (13.8%) [12.2-15.6] | 428 (14.5%) [13.3-15.8] | 26 (8.0%) [5.5-11.5] | 55 (11.3%) [8.8-14.4] | 879 (17.3%) [16.3-18.3] | 1,084 (18.9%) [17.9-19.9] | 468 (18.0%) [16.6-19.5] | 570 (20.1%) [18.6-21.6] |

**Footnote.** CI, confidence interval; NA, counts cannot be displayed due to < 10 in one cell.

[a] expressed in n (%) and 95% confidence interval in brackets; [b] expressed in mean ± standard deviation and 95% confidence interval in brackets; [c] the stay for the delivery was not counted. Differences between intra- groups (across healthcare and assistance professions) were considered statistically significant when there was no overlap between the 95% CIs.

**S5 Table : Stratification by sector of employment (healthcare *versus* social assistance): prevalence of medication dispensed in the year prior to and during pregnancy by occupational groups.**

| | Executives | | | | Intermediate occupations | | | | | | Employees | | | |
|---|---|---|---|---|---|---|---|---|---|---|---|---|---|---|
| | PhysiciansN=2,212 | | Psychologists N=817 | | Nurses N=9,521 | | Social workers N=4,640 | | Rehabilitation professionals N=959 | | Healthcare Assistants N=11,004 | | Hospital service workers N=5,602 | |
| | ***Health-care*** *N=1,914* | ***Social assistance*** *N=47* | ***Health-care*** *N=349* | ***Social assistance*** *N=299* | ***Health-care*** *N=7,819* | ***Social assistance*** *N=936* | ***Health-care*** *N=323* | ***Social assistance*** *N=2,774* | ***Health-care*** *N=452* | ***Social assistance*** *N=277* | ***Health-care*** *N=5,221* | ***Social assistance*** *N=4,355* | ***Health-care*** *N=2,347* | ***Social assistance*** *N=2,917* |
| **DISPENSED MEDICATIONS** | | | | | | | | | | | | | | |
| **Medication dispensed in the year prior to pregnancy** | | | | | | | | | | | | | | |
| **At least one medication** [a] | 1,746 (91.2%) [89.9-92.4] | 45 (95.7%) [85.7-98.8] | 337 (96.6%) [94.1-98.0] | 281 (94.0%) [90.7-96.2] | 7,287 (93.2%) [92.6-93.7] | 870 (92.9%) [91.1-94.4] | 310 (96.0%) [93.2-97.6] | 2,647 (95.4%) [94.6-96.1] | 424 (93.8%) [91.1-95.7] | 262 (94.6%) [91.3-96.7] | 4,978 (95.3%) [94.7-95.9] | 4,149 (95.3%) [94.6-95.9] | 2,203 (93.9%) [92.8-94.8] | 2,726 (93.5%) [92.5-94.3] |
| **Number of different medications** [b] | 8.0±6.5 [7.7-8.3] | 9.7±6.1 [7.9-11.5] | 9.8±7.1 [9.1-10.6] | 9.3±6.3 [8.6-10.0] | 8.5±6.6 [8.4-8.7] | 9.2±6.8 [8.8-9.6] | 10.1±7.0 [9.3-10.9] | 10.8±7.7 [10.5-11.1] | 9.2±7.1 [8.6-9.9] | 9.2±7.1 [8.4-10.1] | 10.9±8.2 [10.6-11.1] | 11.8±8.4 [11.6-12.1] | 12.1±9.0 [11.8-12.5] | 12.1±9.2 [11.8-12.4] |
| **A: Alimentary tract and metabolism** [a] | 955 (49.9%) [47.7-52.1] | 27 (57.4%) [43.3-70.5] | 204 (58.5%) [53.2-63.5] | 174 (58.2%) [52.5-63.6] | 3,830 (49.0%) [47.9-50.1] | 461 (49.3%) [46.1-52.5] | 183 (56.7%) [51.2-62.0] | 1,658 (59.8%) [57.9-61.6] | 239 (52.9%) [48.3-57.4] | 152 (54.9%) [49.0-60.6] | 1,471 (62.7%) [60.7-64.6] | 1,766 (60.5%) [58.8-62.3] | 1,471 (62.7%) [60.7-64.6] | 1,766 (60.5%) [58.8-62.3] |
| **B: Blood and blood forming organs** [a] | 766 (40.0%) [37.8-42.2] | 22 (46.8%) [33.3-60.8] | 133 (38.1%) [33.2-43.3] | 117 (39.1%) [33.8-44.8] | 2,830 (36.2%) [35.1-37.3] | 291 (31.1%) [28.2-34.1] | 126 (39.0%) [33.8-44.4] | 998 (36.0%) [34.2-37.8] | 187 (41.4%) [36.9-46.0] | 114 (41.2%) [35.5-47.0] | 626 (26.7%) [24.9-28.5] | 786 (26.9%) [25.4-28.6] | 626 (26.7%) [24.9-28.5] | 786 (26.9%) [25.4-28.6] |
| **M: Musculo-skeletal system** [a] | 634 (33.1%) [31.1-35.3] | 21 (44.7%) [31.4-58.8] | 146 (41.8%) [36.8-47.1] | 131 (43.8%) [38.3-49.5] | 3,223 (41.2%) [40.1-42.3] | 424 (45.3%) [42.1-48.5] | 149 (46.1%) [40.8-51.6] | 1,440 (51.9%) [50.0-53.8] | 180 (39.8%) [35.4-44.4] | 116 (41.9%) [36.2-47.8] | 1,422 (60.6%) [58.6-62.5] | 1,751 (60.0%) [58.2-61.8] | 1,422 (60.6%) [58.6-62.5] | 1,751 (60.0%) [58.2-61.8] |
| **N: Nervous system** [a] | 1,132 (59.1%) [56.9-61.3] | 31 (66.0%) [51.7-77.8] | 231 (66.2%) [61.1-71.0] | 203 (67.9%) [62.4-72.9] | 4,684 (59.9%) [58.8-61.0] | 611 (65.3%) [62.2-68.3] | 225 (69.7%) [64.4-74.4] | 2,082 (75.1%) [73.4-76.6] | 282 (62.4%) [57.8-66.7] | 180 (65.0%) [59.2-70.4] | 1,803 (76.8%) [75.1-78.5] | 2,274 (78.0%) [76.4-79.4] | 1,803 (76.8%) [75.1-78.5] | 2,274 (78.0%) [76.4-79.4] |
| **R: Respiratory system** [a] | 671 (35.1%) [33.0-37.2] | 20 (42.6%) [29.5-56.7] | 163 (46.7%) [41.5-51.9] | 147 (49.2%) [43.5-54.8] | 3,182 (40.7%) [39.6-41.8] | 424 (45.3%) [42.1-48.5] | 163 (50.5%) [45.0-55.9] | 1,489 (53.7%) [51.8-55.5] | 201 (44.5%) [40.0-49.1] | 129 (46.6%) [40.8-52.5] | 1,250 (53.3%) [51.2-55.3] | 1,539 (52.8%) [50.9-54.6] | 1,250 (53.3%) [51.2-55.3] | 1,539 (52.8%) [50.9-54.6] |
| **Folic acid** [a] (Dispensed in the year prior to pregnancy to 1st trimester of pregnancy) | 957 (50.0%) [47.8-52.2] | 31 (66.0%) [51.7-77.8] | 184 (52.7%) [47.5-57.9] | 141 (47.2%) [41.6-52.8] | 3,542 (45.3%) [44.2-46.4] | 413 (44.1%) [41.0-47.3] | 153 (47.4%) [42.0-52.8] | 1,266 (45.6%) [43.8-47.5] | 244 (54.0%) [49.4-58.5] | 148 (53.4%) [47.5-59.2] | 2,040 (39.1%) [37.8-40.4] | 1,650 (37.9%) [36.5-39.3] | 658 (28.0%) [26.3-29.9] | 930 (31.9%) [30.2-33.6] |
| **Psychotropic medications** [a] | NA | NA | 45 (12.9%) | 41 (13.7%) | 726 (9.3%) | 104 (11.1%) | 47 (14.6%) | 392 (14.1%) | 42 (9.3%) | 32 (11.6%) | 791 (15.2%) | 730 (16.8%) | 424 (18.1%) | 538 (18.4%) |

| | | | | | | | | | | | | | | |
|---|---|---|---|---|---|---|---|---|---|---|---|---|---|---|
| | | | [9.8-16.8] | [10.3-18.1] | [8.7-9.9] | [9.3-13.3] | [11.1-18.8] | [12.9-15.5] | [6.9-12.3] | [8.3-15.9] | [14.2-16.1] | [15.7-17.9] | [16.6-19.7] | [17.1-19.9] |
| **Benzodiazepines** [a] | NA | NA | 35 (10.0%) [7.3-13.6] | 35 (11.7%) [8.5-15.8] | 543 (6.9%) [6.4-7.5] | 80 (8.5%) [6.9-10.5] | 41 (12.7%) [9.5-16.8] | 311 (11.2%) [10.1-12.4] | 28 (6.2%) [4.3-8.8] | 20 (7.2%) [4.7-10.9] | 626 (12.0%) [11.1-12.9] | 576 (13.2%) [12.3-14.3] | 331 (14.1%) [12.8-15.6] | 411 (14.1%) [12.9-15.4] |
| **Antidepressants** [a] | NA | NA | 19 (5.4%) [3.5-8.3] | 15 (5.0%) [3.1-8.1] | 318 (4.1%) [3.7-4.5] | 49 (5.2%) [4.0-6.9] | 22 (6.8%) [4.5-10.1] | 188 (6.8%) [5.9-7.8] | 25 (5.5%) [3.8-8.0] | 10 (3.6%) [2.0-6.5] | 366 (7.0%) [6.3-7.7] | 338 (7.8%) [7.0-8.6] | 184 (7.8%) [6.8-9.0] | 265 (9.1%) [8.1-10.2] |
| **Analgesics** [a] | 1,006 (52.6%) [50.3-54.8] | 29 (61.7%) [47.4-74.2] | 213 (61.0%) [55.8-66.0] | 191 (63.9%) [58.3-69.1] | 4,191 (53.6%) [52.5-54.7] | 551 (58.9%) [55.7-62.0] | 206 (63.8%) [58.4-68.8] | 1,945 (70.1%) [68.4-71.8] | 264 (58.4%) [53.8-62.9] | 166 (59.9%) [54.1-65.5] | 3,552 (68.0%) [66.8-69.3] | 3,202 (73.5%) [72.2-74.8] | 1,702 (72.5%) [70.7-74.3] | 2,166 (74.3%) [72.6-75.8] |
| **Musculoskeletal medications** [a] | 628 (32.8%) [30.7-34.9] | 21 (44.7%) [31.4-58.8] | 145 (41.5%) [36.5-46.8] | 131 (43.8%) [38.3-49.5] | 3,215 (41.1%) [40.0-42.2] | 422 (45.1%) [41.9-48.3] | 148 (45.8%) [40.5-51.3] | 1,438 (51.8%) [50.0-53.7] | 180 (39.8%) [35.4-44.4] | 116 (41.9%) [36.2-47.8] | 2,843 (54.5%) [53.1-55.8] | 2,578 (59.2%) [57.7-60.6] | 1,420 (60.5%) [58.5-62.5] | 1,749 (60.0%) [58.2-61.7] |
| **NSAIDs** [a] | 635 (33.2%) [31.1-35.3] | 20 (42.6%) [29.5-56.7] | 145 (41.5%) [36.5-46.8] | 132 (44.1%) [38.6-49.8] | 3,250 (41.6%) [40.5-42.7] | 431 (46.0%) [42.9-49.2] | 146 (45.2%) [39.9-50.7] | 1,447 (52.2%) [50.3-54.0] | 180 (39.8%) [35.4-44.4] | 118 (42.6%) [36.9-48.5] | 2,858 (54.7%) [53.4-56.1] | 2,569 (59.0%) [57.5-60.4] | 1,412 (60.2%) [58.2-62.1] | 1,734 (59.4%) [57.7-61.2] |
| **Medication dispensed during pregnancy** | | | | | | | | | | | | | | |
| **At least one medication** [a] | 1,796 (93.8%) [92.7-94.8] | 46 (97.9%) [88.9-99.6] | 340 (97.4%) [95.2-98.6] | 288 (96.3%) [93.5-97.9] | 7,377 (94.3%) [93.8-94.8] | 889 (95.0%) [93.4-96.2] | 302 (93.5%) [90.3-95.7] | 2,647 (95.4%) [94.6-96.1] | 437 (96.7%) [94.6-98.0] | 265 (95.7%) [92.6-97.5] | 4,885 (93.6%) [92.9-94.2] | 4,114 (94.5%) [93.8-95.1] | 2,138 (91.1%) [89.9-92.2] | 2,674 (91.7%) [90.6-92.6] |
| **Number of different medications** [b] | 7.0±5.0 [6.8-7.3] | 8.6±4.8 [7.2-10.0] | 8.8±5.7 [8.2-9.4] | 7.8±5.2 [7.2-8.4] | 7.2±4.9 [7.1-7.3] | 7.6±5.2 [7.3-8.0] | 8.3±5.7 [7.7-8.9] | 8.4±5.4 [8.2-8.6] | 7.5±4.6 [7.1-8.0] | 7.8±5.2 [7.2-8.4] | 8.0±5.7 [7.9-8.2] | 8.4±5.8 [8.2-8.6] | 7.9±6.0 [7.7-8.2] | 8.2±6.3 [8.0-8.5] |
| **A: Alimentary tract and metabolism** [a] | 1,519 (79.4%) [77.5-81.1] | 39 (83.0%) [69.9-91.1] | 296 (84.8%) [80.7-88.2] | 244 (81.6%) [76.8-85.6] | 6,179 (79.0%) [78.1-79.9] | 752 (80.3%) [77.7-82.8] | 254 (78.6%) [73.8-82.8] | 2,215 (79.8%) [78.3-81.3] | 366 (81.0%) [77.1-84.3] | 237 (85.6%) [80.9-89.2] | 4,044 (77.5%) [76.3-78.6] | 3,359 (77.1%) [75.9-78.4] | 1,694 (72.2%) [70.3-74.0] | 2,148 (73.6%) [72.0-75.2] |
| **B: Blood and blood forming organs** [a] | 1,231 (64.3%) [62.1-66.4] | 40 (85.1%) [72.3-92.6] | 255 (73.1%) [68.2-77.5] | 206 (68.9%) [63.4-73.9] | 5,193 (66.4%) [65.4-67.5] | 617 (65.9%) [62.8-68.9] | 226 (70.0%) [64.8-74.7] | 1,835 (66.1%) [64.4-67.9] | 310 (68.6%) [64.2-72.7] | 201 (72.6%) [67.0-77.5] | 3,170 (60.7%) [59.4-62.0] | 2,573 (59.1%) [57.6-60.5] | 1,221 (52.0%) [50.0-54.0] | 1,570 (53.8%) [52.0-55.6] |
| **M: Musculo-skeletal system** [a] | NA | NA | 36 (10.3%) [7.5-14.0] | 29 (9.7%) [6.8-13.6] | 718 (9.2%) [8.6-9.8] | 89 (9.5%) [7.8-11.6] | 29 (9.0%) [6.3-12.6] | 388 (14.0%) [12.7-15.3] | 38 (8.4%) [6.2-11.3] | 22 (7.9%) [5.3-11.7] | 807 (15.5%) [14.5-16.5] | 753 (17.3%) [16.2-18.4] | 413 (17.6%) [16.1-19.2] | 536 (18.4%) [17.0-19.8] |
| **N: Nervous system** [a] | 961 (50.2%) [48.0-52.4] | 30 (63.8%) [49.5-76.0] | 213 (61.0%) [55.8-66.0] | 185 (61.9%) [56.2-67.2] | 4,100 (52.4%) [51.3-53.5] | 521 (55.7%) [52.5-58.8] | 208 (64.4%) [59.0-69.4] | 1,828 (65.9%) [64.1-67.6] | 252 (55.8%) [51.1-60.3] | 149 (53.8%) [47.9-59.6] | 3,339 (64.0%) [62.6-65.2] | 3,023 (69.4%) [68.0-70.8] | 1,524 (64.9%) [63.0-66.8] | 1,944 (66.6%) [64.9-68.3] |

| | | | | | | | | | | | | | | |
|---|---|---|---|---|---|---|---|---|---|---|---|---|---|---|
| **R: Respiratory system** [a] | 531 (27.7%) [25.8-29.8] | 18 (38.3%) [25.8-52.6] | 133 (38.1%) [33.2-43.3] | 117 (39.1%) [33.8-44.8] | 2,525 (32.3%) [31.3-33.3] | 326 (34.8%) [31.8-37.9] | 125 (38.7%) [33.6-44.1] | 1,108 (39.9%) [38.1-41.8] | 161 (35.6%) [31.3-40.1] | 100 (36.1%) [30.7-41.9] | 1,919 (36.8%) [35.5-38.1] | 1,757 (40.3%) [38.9-41.8] | 874 (37.2%) [35.3-39.2] | 1,099 (37.7%) [35.9-39.4] |
| **Psychotropic medications** [a] | NA | NA | 15 (4.3%) [2.6-7.0] | 17 (5.7%) [3.6-8.9] | 295 (3.8%) [3.4-4.2] | 44 (4.7%) [3.5-6.3] | 18 (5.6%) [3.6-8.6] | 154 (5.6%) [4.8-6.5] | NA | NA | 312 (6.0%) [5.4-6.7] | 275 (6.3%) [5.6-7.1] | 141 (6.0%) [5.1-7.0] | 203 (7.0%) [6.1-7.9] |
| **Benzodiazepines** [a] | NA | NA | 10 (2.9%) [1.6-5.2] | 11 (3.7%) [2.1-6.5] | 174 (2.2%) [1.9-2.6] | 28 (3.0%) [2.1-4.3] | 12 (3.7%) [2.1-6.4] | 102 (3.7%) [3.0-4.4] | NA | NA | 200 (3.8%) [3.3-4.4] | 182 (4.2%) [3.6-4.8] | 100 (4.3%) [3.5-5.2] | 147 (5.0%) [4.3-5.9] |
| **Antidepressants** [a] | NA | NA | NA | NA | 112 (1.4%) [1.2-1.7] | 17 (1.8%) [1.1-2.9] | NA | NA | NA | NA | 127 (2.4%) [2.0-2.9] | 110 (2.5%) [2.1-3.0] | 57 (2.4%) [1.9-3.1] | 77 (2.6%) [2.1-3.3] |
| **Analgesics** [a] | 888 (46.4%) [44.2-48.6] | 27 (57.4%) [43.3-70.5] | 205 (58.7%) [53.5-63.8] | 175 (58.5%) [52.9-64.0] | 3,855 (49.3%) [48.2-50.4] | 492 (52.6%) [49.4-55.7] | 197 (61.0%) [55.6-66.2] | 1,735 (62.5%) [60.7-64.3] | 243 (53.8%) [49.2-58.3] | 143 (51.6%) [45.8-57.4] | 3,162 (60.6%) [59.2-61.9] | 2,900 (66.6%) [65.2-68.0] | 1,459 (62.2%) [60.2-64.1] | 1,867 (64.0%) [62.2-65.7] |
| **Musculoskeletal medications** [a] | NA | NA | 35 (10.0%) [7.3-13.6] | 28 (9.4%) [6.6-13.2] | 710 (9.1%) [8.5-9.7] | 88 (9.4%) [7.7-11.4] | 29 (9.0%) [6.3-12.6] | 384 (13.8%) [12.6-15.2] | 37 (8.2%) [6.0-11.1] | 22 (7.9%) [5.3-11.7] | 798 (15.3%) [14.3-16.3] | 746 (17.1%) [16.0-18.3] | 411 (17.5%) [16.0-19.1] | 535 (18.3%) [17.0-19.8] |
| **NSAIDs** [a] | NA | NA | 47 (13.5%) [10.3-17.4] | 33 (11.0%) [8.0-15.1] | 891 (11.4%) [10.7-12.1] | 102 (10.9%) [9.1-13.1] | 34 (10.5%) [7.6-14.3] | 433 (15.6%) [14.3-17.0] | 45 (10.0%) [7.5-13.1] | 25 (9.0%) [6.2-13.0] | 938 (18.0%) [16.9-19.0] | 807 (18.5%) [17.4-19.7] | 431 (18.4%) [16.8-20.0] | 565 (19.4%) [18.0-20.8] |

**Footnote.** CI, confidence interval; NA, counts cannot be displayed due to < 10 in one cell.

[a] expressed in n (%) and 95% confidence interval in brackets; [b] expressed in mean ± standard deviation and 95% confidence interval in brackets; [c] the stay for the delivery was not counted. Differences between intra-groups (across healthcare and assistance professions) were considered statistically significant when there was no overlap between the 95% CIs.

**S6 Table : Prevalence of specific medication dispensed in the year prior to and during pregnancy by occupational groups.**

| Specific medication | Executives | | | Intermediate occupations | | | | Employees | | | **Other** N=4,105 |
|---|---|---|---|---|---|---|---|---|---|---|---|
| | Non-healthcare sectors N=24,613 | PhysiciansN= 2,212 | Psychologists N=817 | Non-healthcare sectors N=32,828 | Social workers N=4,640 | Rehabilitation professionals N=959 | Nurses N=9,521 | Non-healthcare sectors N= 106,943 | Healthcare assistants N=11,004 | Hospital service workers N=5,602 | |
| **Folic acid** | | | | | | | | | | | |
| Dispensed in the year prior to pregnancy to 1$^{st}$ trimester of pregnancy | 11,756 (47.8%) [47.1-48.4] | 1,122 (50.7%) [48.6-52.8] | 412 (50.4%) [47.0-53.8] | 13,775 (42.0%) [41.4-42.5] | 2,133 (46.0%) [44.5-47.4] | 512 (53.4%) [50.2-56.5] | 4,272 (44.9%) [43.9-45.9] | 36,113 (33.8%) [33.5-34.1] | 4,251 (38.6%) [37.7-39.5] | 1,700 (30.3%) [29.2-31.6] | 1,798 (43.8%) [42.3-45.3] |
| **Psychotropic medications** | | | | | | | | | | | |
| Dispensed in the year prior to pregnancy | 2,758 (11.2%) [10.8-11.6] | 246 (11.1%) [9.9-12.5] | 104 (12.7%) [10.6-15.2] | 4,296 (13.1%) [12.7-13.5] | 647 (13.9%) [13.0-15.0] | 91 (9.5%) [7.8-11.5] | 928 (9.7%) [9.2-10.4] | 15,431 (14.4%) [14.2-14.6] | 1,726 (15.7%) [15.0-16.4] | 1,029 (18.4%) [17.4-19.4] | 464 (11.3%) [10.4-12.3] |
| Dispensed during pregnancy | 1,110 (4.5%) [4.3-4.8] | 105 (4.7%) [3.9-5.7] | 41 (5.0%) [3.7-6.7] | 1,689 (5.1%) [4.9-5.4] | 259 (5.6%) [5.0-6.3] | 30 (3.1%) [2.2-4.4] | 373 (3.9%) [3.5-4.3] | 5,885 (5.5%) [5.4-5.6] | 677 (6.2%) [5.7-6.6] | 369 (6.6%) [6.0-7.3] | 179 (4.4%) [3.8-5.0] |
| **Benzodiazepines** | | | | | | | | | | | |
| Dispensed in the year prior to pregnancy | 2,112 (8.6%) [8.2-8.9] | 191 (8.6%) [7.5-9.9] | 83 (10.2%) [8.3-12.4] | 3,335 (10.2%) [9.8-10.5] | 499 (10.8%) [9.9-11.7] | 63 (6.6%) [5.2-8.3] | 701 (7.4%) [6.9-7.9] | 12,247 (11.5%) [11.3-11.6] | 1,370 (12.5%) [11.8-13.1] | 790 (14.1%) [13.2-15.0] | 361 (8.8%) [8.0-9.7] |
| Dispensed during pregnancy | 665 (2.7%) [2.5-2.9] | 76 (3.4%) [2.8-4.3] | 28 (3.4%) [2.4-4.9] | 1,073 (3.3%) [3.1-3.5] | 158 (3.4%) [2.9-4.0] | 13 (1.4%) [0.8-2.3] | 219 (2.3%) [2.0-2.6] | 3,904 (3.7%) [3.5-3.8] | 432 (3.9%) [3.6-4.3] | 262 (4.7%) [4.2-5.3] | 114 (2.8%) [2.3-3.3] |
| **Antidepressants** | | | | | | | | | | | |
| Dispensed in the year prior to pregnancy | 1,263 (5.1%) [4.9-5.4] | 103 (4.7%) [3.9-5.6] | 44 (5.4%) [4.0-7.2] | 1,984 (6.0%) [5.8-6.3] | 311 (6.7%) [6.0-7.5] | 41 (4.3%) [3.2-5.7] | 410 (4.3%) [3.9-4.7] | 7,029 (6.6%) [6.4-6.7] | 799 (7.3%) [6.8-7.8] | 493 (8.8%) [8.1-9.6] | 214 (5.2%) [4.6-5.9] |
| Dispensed during pregnancy | 530 (2.2%) [2.0-2.3] | 35 (1.6%) [1.1-2.2] | 20 (2.4%) [1.6-3.8] | 765 (2.3%) [2.2-2.5] | 125 (2.7%) [2.3-3.2] | 13 (1.4%) [0.8-2.3] | 147 (1.5%) [1.3-1.8] | 2,465 (2.3%) [2.2-2.4] | 278 (2.5%) [2.2-2.8] | 151 (2.7%) [2.3-3.2] | 80 (1.9%) [1.6-2.4] |
| **Analgesics** | | | | | | | | | | | |
| Dispensed in the year prior to pregnancy | 15,039 (61.1%) [60.5-61.7] | 1,186 (53.6%) [51.5-55.7] | 511 (62.5%) [59.2-65.8] | 22,010 (67.0%) [66.5-67.6] | 3,205 (69.1%) [67.7-70.4] | 557 (58.1%) [54.9-61.2] | 5,184 (54.4%) [53.4-55.4] | 73,188 (68.4%) [68.2-68.7] | 7,739 (70.3%) [69.5-71.2] | 4,102 (73.2%) [72.0-74.4] | 2,412 (58.8%) [57.2-60.3] |
| Dispensed during pregnancy | 13,662 (55.5%) [54.9-56.1] | 1,049 (47.4%) [45.3-49.5] | 480 (58.8%) [55.3-62.1] | 19,994 (60.9%) [60.4-61.4] | 2,846 (61.3%) [59.9-62.7] | 497 (51.8%) [48.7-55.0] | 4,758 (50.0%) [49.0-51.0] | 66,131 (61.8%) [61.5-62.1] | 6,965 (63.3%) [62.4-64.2] | 3,545 (63.3%) [62.0-64.5] | 2,384 (58.1%) [56.6-59.6] |

| | | | | | | | | | | | |
|---|---|---|---|---|---|---|---|---|---|---|---|
| **Musculoskeletal medications** | | | | | | | | | | | |
| Dispensed in the year prior to pregnancy | 10,811 (43.9%) [43.3-44.5] | 740 (33.5%) [31.5-35.4] | 353 (43.2%) [39.8-46.6] | 16,445 (50.1%) [49.6-50.6] | 2,368 (51.0%) [49.6-52.5] | 394 (41.1%) [38.0-44.2] | 3,975 (41.7%) [40.8-42.7] | 56,745 (53.1%) [52.8-53.4] | 6,187 (56.2%) [55.3-57.1] | 3,352 (59.8%) [58.5-61.1] | 2,008 (48.9%) [47.4-50.4] |
| Dispensed during pregnancy | 2,511 (10.2%) [9.8-10.6] | 227 (10.3%) [9.1-11.6] | 82 (10.0%) [8.2-12.3] | 4,258 (13.0%) [12.6-13.3] | 584 (12.6%) [11.7-13.6] | 80 (8.3%) [6.8-10.3] | 887 (9.3%) [8.7-9.9] | 17,296 (16.2%) [16.0-16.4] | 1,751 (15.9%) [15.2-16.6] | 1,008 (18.0%) [17.0-19.0] | 517 (12.6%) [11.6-13.6] |
| **NSAIDs** | | | | | | | | | | | |
| Dispensed in the year prior to pregnancy | 11,062 (44.9%) [44.3-45.6] | 753 (34.0%) [32.1-36.0] | 353 (43.2%) [39.8-46.6] | 16,595 (50.6%) [50.0-51.1] | 2,369 (51.1%) [49.6-52.5] | 395 (41.2%) [38.1-44.3] | 4,017 (42.2%) [41.2-43.2] | 56,834 (53.1%) [52.8-53.4] | 6,199 (56.3%) [55.4-57.3] | 3,330 (59.4%) [58.2-60.7] | 2,024 (49.3%) [47.8-50.8] |
| Dispensed during pregnancy | 3,195 (13.0%) [12.6-13.4] | 323 (14.6%) [13.2-16.1] | 103 (12.6%) [10.5-15.1] | 4,939 (15.0%) [14.7-15.4] | 666 (14.4%) [13.4-15.4] | 96 (10.0%) [8.3-12.1] | 1,103 (11.6%) [11.0-12.2] | 18,978 (17.7%) [17.5-18.0] | 1,988 (18.1%) [17.4-18.8] | 1,063 (19.0%) [18.0-20.0] | 613 (14.9%) [13.9-16.1] |

**Footnote.** CI, confidence interval; Differences between intra- and inter- groups (across healthcare and assistance professions and non-healthcare professions) were considered statistically significant when there was no overlap between the 95% CIs. Expressed in n (%) and 95% confidence interval in brackets
